# System-wide hematopoietic and immune signaling aberrations in COVID-19 revealed by deep proteome and phosphoproteome analysis

**DOI:** 10.1101/2021.03.19.21253675

**Authors:** Tomonori Kaneko, Sally Esmail, Courtney Voss, Claudio M. Martin, Marat Slessarev, Owen Hovey, Xuguang Liu, Mingliang Ye, Sung Kim, Douglas Fraser, Shawn SC Li

## Abstract

The COVID-19 pandemic, caused by the SARS-CoV-2 virus, has become a global crisis. To gain systems-level insights into its pathogenesis, we compared the blood proteome and phosphoproteome of ICU patients with or without SARS-CoV-2 infection, and healthy control subjects by quantitative mass spectrometry. We find that COVID-19 is marked with hyperactive T cell and B cell signaling, compromised innate immune response, and dysregulated inflammation, coagulation, metabolism, RNA splicing, transcription and translation pathways. SARS-CoV-2 infection causes global reprogramming of the kinome and kinase-substrate network, resulting in defective antiviral defense via the CK2-OPN-IL-12/IFN-α/β axis, lymphocyte cell death via aberrant JAK/STAT signaling, and inactivation of innate immune cells via inhibitory SIRPA, SIGLEC and SLAM family receptor signaling. Our work identifies CK2, SYK, JAK3, TYK2 and IL-12 as potential targets for immunomodulatory treatment of severe COVID-19 and provides a valuable approach and resource for deciphering the mechanism of pathogen-host interactions.

## INTRODUCTION

The severe acute respiratory syndrome coronavirus-2 or SARS-CoV-2, first identified in humans in December 2019^1^, has infected tens of millions of people and claimed nearly 2 million lives worldwide to date. While the majority of individuals infected by the new coronavirus have mild symptoms or are asymptomatic, approximately 10-15% of patients develop severe diseases that require hospitalization. Patients with severe symptoms usually develop acute respiratory distress syndrome (ARDS) ^2^, a major cause of morbidity and mortality. The coronavirus disease 2019 (COVID-19) pandemic has sparked an unprecedented effort from the scientific community to understand the disease mechanism and develop therapeutic and immunization strategies to combat the pandemic, cumulating in the recent approval of two vaccines for emergency use by the US Federal Drug Administration.

Despite these encouraging developments, the biological and biochemical underpinnings of ARDS and other symptoms of severe COVID-19 are not fully understood, which has hampered the development of effective treatment. Studies to date have shown that the SARS-CoV-2 infection elicits a wide array of biochemical and cellular abnormalities that are rooted in an unbalanced immune response to the virus. For example, while cytokines play an important role in antiviral immunity, rapid production of a large amount of proinflammatory cytokines, referred to as the cytokine release syndrome (CRS), is associated with severe COVID-19 cases^3^. Furthermore, severe diseases are characterized frequently with lymphopenia, manifesting in reduced circulating T cells, B cells or/and natural killer (NK) cells ^4^. However, immune profiling has revealed activation of subsets of T cells, extrafollicular B cells and production of neutralizing antibodies in severe COVID-19, suggesting robust cellular and humoral immune responses in these patients ^5,6^. Nevertheless, a large proportion of patients with a strong, early antibody response do poorly in controlling the infection and ultimately succumb to the disease^6^. These dichotomies highlight voids in our knowledge on the fundamental immunological processes perturbed by the SARS-CoV-2 virus.

To decipher the molecular, cellular and immunological basis of COVID-19 in a systematic and unbiased manner, we employed mass spectrometry (MS) and complementary biochemical assays to characterize the peripheral blood – the barometer of the immune system. By comparing the blood proteome and phosphoproteome of the patients under intensive care who tested positive or negative for the SARS-CoV-2 virus with age- and sex-matched healthy controls, we identified the protein/phosphoprotein signatures and the regulatory/signaling pathways associated with severe COVID-19. We show that the SARS-CoV-2^+^ ICU patients are marked with aberrant changes in a wide range of hematopoietic and immunological processes and pathways. Combined with comprehensive cytokine/chemokine and antibody profiling, our MS analysis not only captured the major defects identified to date for COVID-19, but also generated novel insights into the signaling and regulatory mechanisms underlying the aberrant immune responses mediated by T cells, B cells, NK cells and myeloid cells. Importantly, our work has identified numerous potential therapeutic targets, including the casein kinase 2 (CK2), the spleen tyrosine kinase (SYK), the Janus kinases JAK3 and TYK2, and interleukin-12 (IL-12), for the development of targeted immunomodulatory therapies for the treatment of patients with severe COVID-19 diseases.

## RESULTS

### Quantitative MS analysis of the blood reveals distinctive features of the COVID-19 proteome

We employed mass spectrometry (MS) to identify proteins and phosphoproteins in the blood of critically ill COVID-19 patients in comparison to age-and sex-matched critically ill SARS-CoV-2 negative and healthy control subjects. Specifically, the peripheral blood mononuclear cells (PBMCs) were isolated, respectively, from the blood of 5 ICU patients who tested positive for the SARS-CoV-2 RNA (the COV group; median years of age= 61.0; IQR = 54.8–67.0), 5 SARS-CoV-2-negative ICU patients (the ICU group; median years of age = 58.0; IQR = 52.5–63.0), and 5 healthy individuals (the HC group; median years of age = 57.5; IQR = 52.8–62.8) (Table S1). To gauge the proteome and phosphoproteome dynamics accompanying disease progression, we included in the MS analysis serial blood samples from the COV group drawn on days 1, 7 and 10 (or D1, D7, and D10) of ICU admission. Peptides from the 25 PBMC were isobarically labeled with tandem mass tags (TMT) in three batches (Table S2) and subjected to liquid chromatography (LC)-MS/MS analysis (Fig. S1, Table S3). The phosphoproteome identification was facilitated by SH2 superbinder enrichment for the pTyr-containing peptides ^7^ and by IMAC (immobilized metal affinity chromatography) enrichment for the pSer/pThr-containing peptides (Fig. S1). The MS analysis identified 3,236 non-redundant proteins containing 2,317 Ser/Thr and 394 Tyr unique phosphorylation sites (Tables S4-6).

Clustering of the 894 proteins that differed significantly in abundance correctly identified the COV, ICU and HC cohorts, suggesting distinctive protein expression patterns in the different PBMCs (Fig. 1A). In general, proteins over-expressed in the patients were found under-expressed in the healthy controls and vice versa (i.e., Clusters 1 and 5, Fig. 1A). In contrast, fewer than 130 proteins (belonging to the clusters 2, 3 and 4, Fig. 1A) were significantly different in expression between COV and ICU, suggesting that the proteomes of the two patient groups are similar. Gene ontology (GO) analysis revealed a number of significantly enriched processes that included exocytosis, innate and adaptive immune responses, interleukin signaling, platelet activation, redox response, and glycolysis (Fig. 1B). Intriguingly, interleukin-12 (IL-12) signaling emerged as a significant feature. As described later, IL-12 expression was drastically reduced in the COV PBMCs. Because functionally related proteins tend to interact with one another, we employed the protein-protein interaction (PPI) clustering feature of Metascape to identify the enriched PPI networks for the 521 differentially expressed proteins between COV and HC (Fig. S2). This led to the identification of four PPI networks enriched in the COVID-19 samples with functions in cell cycle phase transition^8^, RNA processing, catabolism ^9^ and exocytosis and stress responses (Fig. S3). The PPI network for RNA processing comprises a spliceosome and a translation regulation subnetwork (Fig. 1C), reinforcing the previous finding that SARS-CoV-2 suppresses global mRNA splicing and translation^10^.

**Fig. 1.**
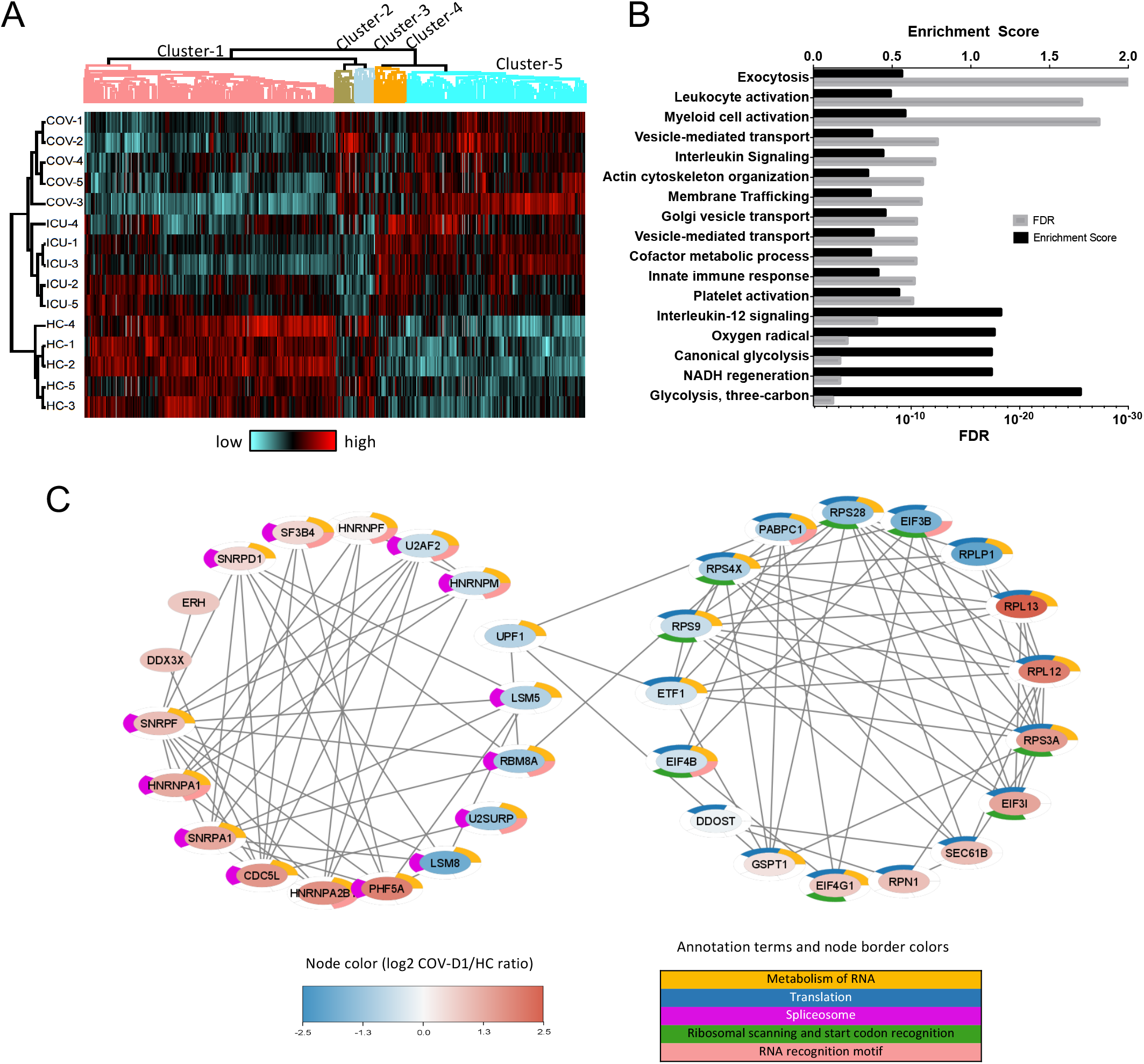
Proteome signatures of COVID-19. (**A**) Heatmap representation of differentially expressed proteins between the proteome of the COV (n=5, day 1 samples or COV-D1), ICU (n=5, day 1) and HC (n=5) PBMCs ordered by hierarchical clustering of the normalized intensities (z-scores) of the 894 proteins that were significantly distinguished by an ANOVA test between the groups. Over-expressed proteins (compared to the mean for each protein) are shown in red, and under-expressed proteins are shown in cyan. (**B**) Gene Ontology (GO) enrichment of the differentially expressed proteins based on analysis with Metascape. Significant features intersecting each GO category were compared to the background dataset as an enrichment score and FDR (Benjamini-Hochberg) q-value from Fisher’s exact test. (**C**) A PPI network for RNA splicing (left) and translation (right) identified from the significantly regulated proteins (constructed using STRING). The node color gradient corresponds to the log2 COV-D1/HC ratio. The node border is colored with the five major functional annotation terms enriched in the network.

### Systematic changes in the immune response, coagulation, platelet activation and metabolic pathways in COVID-19

To gain further insights into the cellular and biochemical defects caused by SARS-CoV-2, we next focused on the differentially expressed proteins in specific pathways, taking advantage of the quantitative proteome data. In keeping with an essential role for regulated exocytosis (Fig. 1B) in the degranulation and activation of myeloid cells, we found that the neutrophil activation and monocyte/macrophage differentiation pathways^11^ were significantly upregulated in the COV group (D1, D7 and D10) compared to HC or ICU. Phagocytosis/endocytosis, which plays an important part in the elimination of virus-infected cells, was also significantly elevated in COV. Specifically, the Fc receptor-dependent phagocytosis pathway was progressively upregulated from D1 to D7 and D10 for the COV group, coinciding with increased antibody response to SARS-CoV-2 (*vide infra*). In agreement with the GO enrichment analysis (Fig. S4), the complement and coagulation pathway, which plays key roles in innate immunity and hemostasis in an interactive manner, was significantly elevated in COV in a progressive manner from D1 to D10, but not in ICU. The same was observed for platelet activation. Indeed, we found that the fibrinogens FGA, FGB and FGG, which when cleaved by thrombin, form fibrin-based blood clots, were significantly upregulated. Similarly, expression of the complement regulators such as C1QA, C3, C5, C7, CFB, CFD, CFH, and CFHR5 was significantly increased in COV (Table S4). These findings are consistent with the results from published data showing that significant upregulation of cellular proteins related to neutrophil activation and blood coagulation^12,13^ and the activation of the complement and platelet pathways ^9,14,15^ in COVID-19.

Overproduction of proinflammatory cytokines is a hallmark of severe SARS-COV-2 infection. Indeed, we found that the cytokine/chemokine signaling pathway significantly upregulated in the COV, but not the ICU group. Because antigen processing and presentation is a significantly enriched process in the GO analysis (Fig. S4B), we analyzed the MHC-I and MHC-II mediated antigen presentation pathways and found that the former significantly up-regulated and the latter down-regulated in COV (Fig. 2), suggesting defects in antigen presentation by professional antigen presenting cells^11^. Massive metabolic change in the host cell is another hallmark of SARS-CoV-2 infection^9,16^. We found, indeed, that the TCA cycle was significantly downregulated while glycolysis significantly upregulated in COV, and the trend became more pronounced with disease progression. This suggests the SARS-CoV-2 infection reprogrammed energy production from aerobic metabolism inside the mitochondria to anaerobic metabolism outside mitochondria. In agreement with a wholesale change in metabolism, both the nucleic acid metabolism (not shown) and fatty acid synthesis and metabolism pathways (Fig. 2) were found significantly increased in COV.

**Fig. 2.**
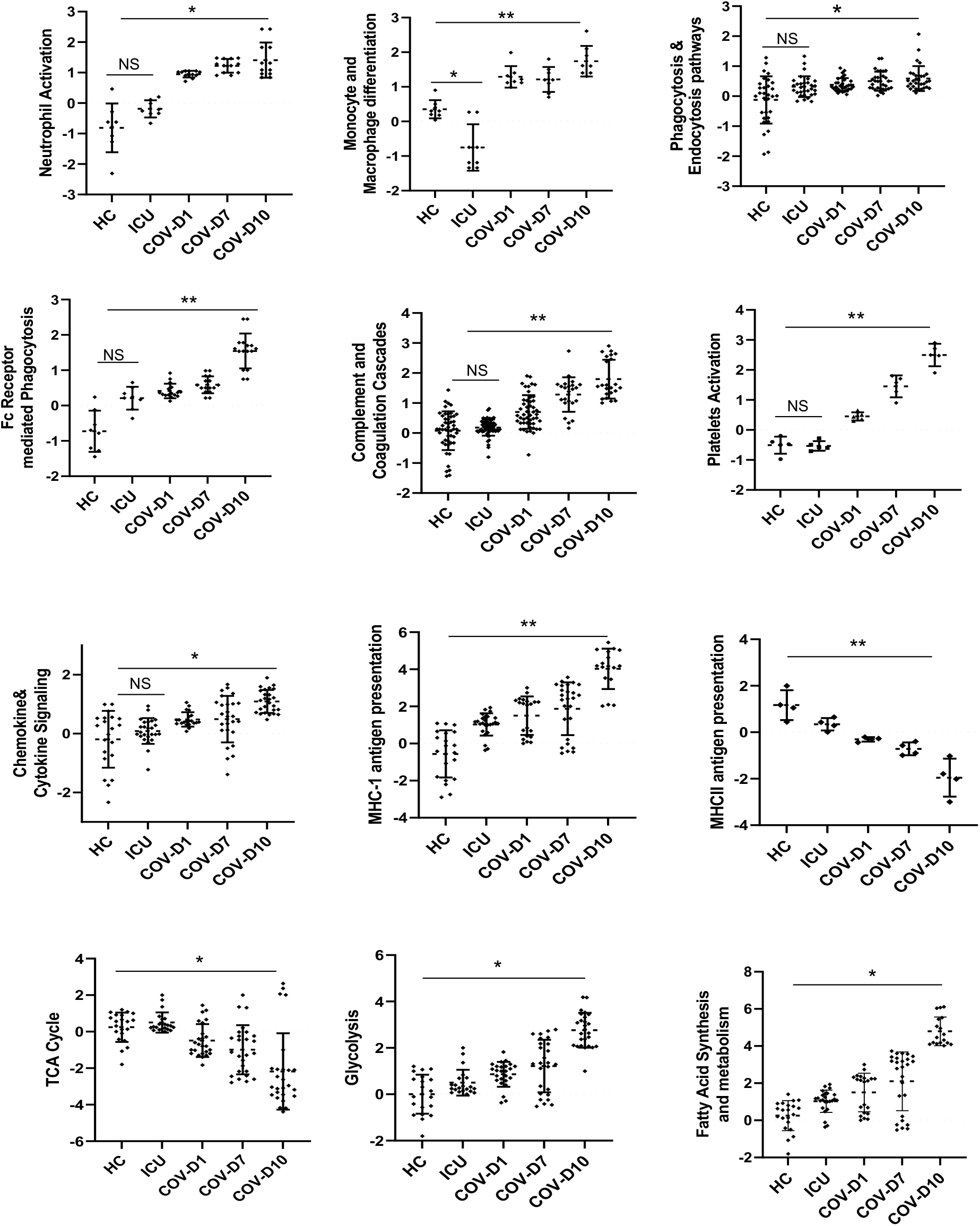
Pathways and processes perturbed in the PBMCs of critically ill COVID-19 patients. Each spot corresponds to a protein within the specified pathway based on GO annotation. The y-axis shows the log2 value of the fold change in protein expression for the group (relative to the mean of the whole proteome). The p values were calculated based on ANOVA test, *p<0.05, **p<0.002, NS, not significant.

### Kinome reprogramming by SARS-CoV-2

Phosphorylation plays a pivotal role in cellular signal transduction and communication of the cell with its environment. The widespread changes in the cellular and immunological pathways implies that differential protein phosphorylation may play a role. Indeed, of the ∼3,000 Ser/Thr and Tyr phosphosites identified, 25% (670 pS/pT and 94 pY sites) differed significantly between COV, ICU and HC (Tables S5 and S6). The proteins with differentially Ser/Thr phosphorylation were found enriched in various metabolic processes, RNA processing or transcription, virus-host or cell-cell interaction, and protein folding (Fig. S5). An increase in misfolded protein response suggests virus infection-induced ER stress is an important feature of COVID-19 ^17^. In contrast, the proteins with differential Tyr phosphorylation were involved mainly in adaptive and immune responses (Fig. S6). This is expected given the importance of Tyr phosphorylation in the proximal signaling by immunoreceptors and their associated proteins.

To understand the role of protein kinases in the immune response to SARS-CoV-2 infection, we next focused on characterizing kinase phosphorylation and activation. The activity of a kinase is often regulated/induced by the phosphorylation of specific residues, some of which are located in the activation loop. Therefore, the phosphorylation status of the regulatory site(s) provides a facile readout of the activity of the corresponding kinase^7^. Based on this rationale, seven of the dozen detected tyrosine kinases (TKs), including LYN, SYK, BTK, HCK, FER, TEC and PTK2, were more active in COV than HC (Fig. 3A). It is important to note that LYN, SYK and BTK, which play pivotal roles in the signal transduction by the B cell receptor (BCR) and a variety of other immunoreceptors, were more activated in COV than ICU. LYN was hyper-phosphorylated on multiple Tyr or Ser sites, including Y473 within the kinase domain and Y397 in the activation loop (which is identical in sequence to the HCK activation loop). In contrast, the phosphorylation of the inhibitory Y194 residue ^18^ was significantly reduced (Fig. 3A). The phosphorylation profile of LYN together with its increased expression (Fig. S7), suggests that SARS-CoV-2 caused a significant increase in LYN activity. Similarly, both the activity (i.e, activation loop Tyr525 phosphorylation) and the expression of SYK were significantly increased in COV vs. ICU or HC (Fig. 3A, Fig. S7B). BTK and TEC, both playing a critical role in PLCγ1 activation and Ca^2+^ mobilization downstream of antigen receptors ^19^, were also highly activated in the COV group. Together, these data suggest that B cell signaling is escalated significantly in the COVID-19 patients. In contrast to the B cell kinases, the status of the T cell receptor (TCR) proximal kinase LCK was less clear. LCK expression was drastically reduced in the COV PBMCs (Fig. S7A), consistent with pronounced T cell lymphopenia in severe COVID-19 cases. However, phosphorylation of the inhibitory Y505 residue was significantly decreased in COV. Moreover, CSK, the kinase that phosphorylates Y505, was reduced in both phosphorylation and expression (Fig. 3A; Fig. S7). Therefore, it is likely that LCK was more active in the COVID-19 patients despite reduced expression.

**Fig. 3.**
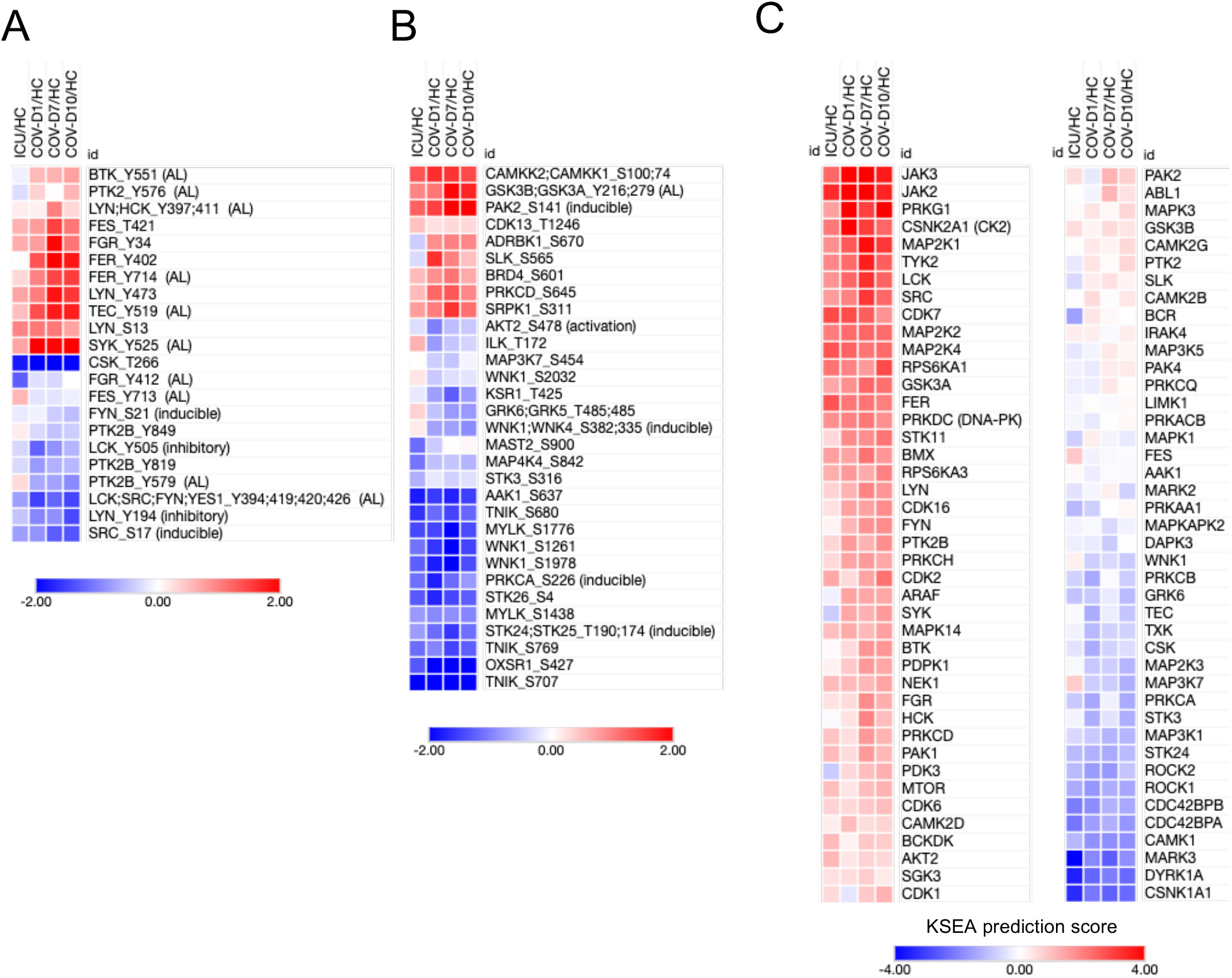
Kinase phosphorylation and activity status in the COV and ICU PBMCs. The averaged log2 ratio of matched samples (e.g., the mean value of ICU1/HC1, ICU2/HC2…ICU5/HC5) are displayed as a heatmap with clustering. (**A**) Detected tyrosine kinases (TKs). (**B**) Ser/Thr kinases (STKs) showing significant changes in phosphorylation between patients and the heathy controls. See Supplementary Table S3 for a complete list of all detected kinase phosphorylation sites. “AL”: kinase activation loop, “inducible”: the phosphorylation induces kinase activity, “inhibitory”: the phosphorylation inhibits kinase activity. The annotations are based on PhosphositePlus, except for LCK-pY194 and AKT2 pSer478. (C) Kinase activity predicted using Kinase-Substrate Enrichment Analysis (KSEA). The combined phosphoproteome data (pTyr and IMAC datasets) were used to analyze which kinases may be activated or supressed in the COVID-19 patients compared to healthy controls, based on enrichment of phosphorylated substrates in the COV samples. The prediction used both the PhosphositePlus and NetworKIN datasets. The list of kinases was filtered and the kinases detected in any of the mass spec injections were retained in the figure.

Numerous Ser/Thr kinases (STKs) showed significant changes in phosphorylation (Fig. 3B). The phosphorylation of 9 STKs, including on activation or inducible residues, was significantly increased, suggesting potential activation of these kinase. Of note, SLK and PRKCD/PKCδ, which can mediate/promote apoptosis, were significantly over-phosphorylated whereas AKT2, critical for cell survival, is under-phosphorylated at its activation site residue S478 ^20^, suggesting that these kinases may work in concert to promote PBMC apoptosis and thereby, contributing to lymphopenia for the COVID-19 patients (Table S1). The β-adrenergic receptor kinase ADRBK1/GRK2, a hallmark of cardiac stress and heart failure^21^, was highly and selectively phosphorylated in the COV PBMCs, suggesting that GRK2 may contribute to cardiac dysfunction associated with COVID-19 ^22^. In support of this possibility, elevated myocardial and lymphocyte GRK2 expression and activation has been associated with heart failure^23^.

In contrary to the activated kinases, WNK1 exhibited significantly reduced phosphorylation on multiple sites, including the activation loop S382 residue, suggesting that WNK1 activity is inhibited in the COVID-19 patients. As an important regulator of blood pressure and electrolyte homeostasis^24^, WNK1 may play a role in regulating blood pressure in COVID-19 patients. The TRAF2- and NCK-interacting kinase (TNIK) is another kinase with reduced phosphorylation on multiple sites. TNIK is an essential activator of the Wnt signaling pathway, and the reduced TNIK activation together with increased activation of GSK3B/3A ^25^, may contribute collectively to reduced Wnt signaling. Because Wnt signaling is involved in dendritic cell (DC) maturation and survival of regulatory T cells^26^, the aberrant inactivation of TNIK and activation of GSK3B/3A may underly the reduced Treg cell population found in hospitalized patients ^27^. Moreover, TNIK is required for canonical NF-κB signaling, and reduced TNIK activity may contribute to reduced antiviral response^28^.

While the MS analysis yielded quantitative phosphorylation information for numerous kinases (Table S7), not all expressed kinases were detected for phosphorylation due to the stochastic nature of mass spectrometry and the unfavorable chromatographic behavior for some phosphopeptides. However, the activity of the kinases missed in the phosphoproteomic analysis may be inferred by Kinase-Substrate Enrichment Analysis (KSEA). Indeed, KSEA predicted 55 kinases with increased activity and 25 with decreased activity in COV compared to HC (Fig. 3C). Moreover, the vast majority of kinases were predicted to be more active in COV than ICU, suggesting that the SARS-CoV-2 infection reprogrammed the kinome, rendering it more active.

Consistent with enhanced cytokine signaling (Fig. 2), the most activated kinases based on KSEA are the Janus kinases JAK3, JAK2, and TYK2. Because deficiency in either *JAK3* or *TYK2* causes immunodeficiency, their over-activation may underly the autoimmune condition associated with severe COVID-19 ^29^. Intriguingly, PRKG1, a key regulator of nitric oxide (NO)/cGMP signaling, and CK2, a promiscuous STK, were also highly active in COV compared to HC. CK2 was identified in a recent study as the top kinase activated in the Vero E6 cells following SARS-CoV-2 infection ^8^. Our data, therefore, further underscores a critical role for CK2 in COVID-19 pathogenesis. The placement of LCK as the 6^th^ most active kinase based on KSEA reinforces the notion that T cell receptor (TCR) signaling is activated in the COVID-19 patients. It is also worth noting that PRKDC/DNA-PK was strongly activated in COV. Because DNA-PK deficiency has been shown to potentiates cGAS-mediated antiviral innate immunity^30^, the increased DNA-PK activity suggests inhibition of antiviral immune response.

Several kinases, including DYRK1A, CSNK1A1 (CK1α) and MARK3, were inactivated in COV based on KSEA, suggesting defects in alternative RNA splicing, Wnt signaling, and microtube dynamics. The oppositive activity patterns of CK2 and CK1 may be due to their differential expression (Table S4). Collectively, these data suggests that the SARS-CoV-2 infection causes systematic reprogramming of the active kinome and the associated processes or pathways. While the signaling pathways for BCR, TCR, cytokine and cardiac stress were significantly activated in the COVID-19 PBMCs, others, including Wnt signaling, RNA splicing and blood pressure regulation, were apparently suppressed.

### Rewiring of the protein kinase-substrate network by SARS-CoV-2

The emergence of CK2 as a highly activated kinase based on substrate enrichment is intriguing. Unlike many other kinases, the activity of CK2 is not regulated by the activation loop, but by the tetramerization of catalytic (α or α’) and regulatory β subunits into the holoenzyme (α/α’)_2_β_2_. However, the catalytic subunit may exhibit activity independently of the regulatory subunit ^31^. Although we were not able to detect CK2α phosphorylation, our phosphoproteome dataset contained 44 CK2 substrates that were differentially phosphorylated between COV and HC (Table S8), providing valuable information on the cellular processes/pathways regulated by CK2 in the context of COVID-19. In agreement with the known functions of CK2, the CK2 substrate network is enriched with proteins involved in DNA binding, RNA processing/transcription, protein translation and folding. Of note, both the DNA replication licensing factor MCM2 (eg., pS27 and pS139, activating) and the RNA polymerase-associated protein LEO1 were phosphorylated on multiple sites. Similarly, the transcriptional activator PURB, translation initiation factor EIF3 and several ribonuclear proteins were highly phosphorylated in the COV group. Related to this, we found the heat shock protein HSP-90 phosphorylated at the activation residue S226, implicating increased demand for protein folding and/or ER stress.

Besides proteins involved in the DNA-RNA-protein genetic information cascade, the most highly phosphorylated (in both number of sites and degree of phosphorylation) CK2 substrate is osteopontin (OPN or SPP1) (Fig. 4A; Fig. S8). Because phosphorylation, in general, inhibits osteopontin function, it is likely that CK2 shut down the many osteopontin signaling pathways through phosphorylation. OPN plays a critical role in the crosstalk between innate and adaptive immunity by regulating the differentiation of Th1 and Th17 cells, the production of IL-12 by macrophages and type I interferon response by plasmacytoid dendritic cells (pDCs) ^32,33^. Thus, the CK2-OPN-IL-12/IFN-α/β axis may underly a multitude of immunological defects associated with severe SARS-CoV-2 infection, including under-representation of Th1 subset in virus reactive CD4+ T cells^27^ and impaired type I interferon response^34,35^. The effect of IL-12 on the development of Th1 may be aided by CD44, which plays an essential role in the survival and memory development in Th1 cells^36^. We found significantly reduced in CD44-S706 phosphorylation in COV, suggesting reduced activation and signaling through CD44. Moreover, intracellular OPN enhances TLR7 and TLR9 (key antiviral innate immune receptor) and Tfh/Bcl6+ cells. SARS-CoV-2 infection prevents Tfh (Bcl6+) cell generation and durable antibody production^37^. Therefore, the CK2-OPN axis could cause defects in inducing strong anti-viral responses.

**Fig. 4.**
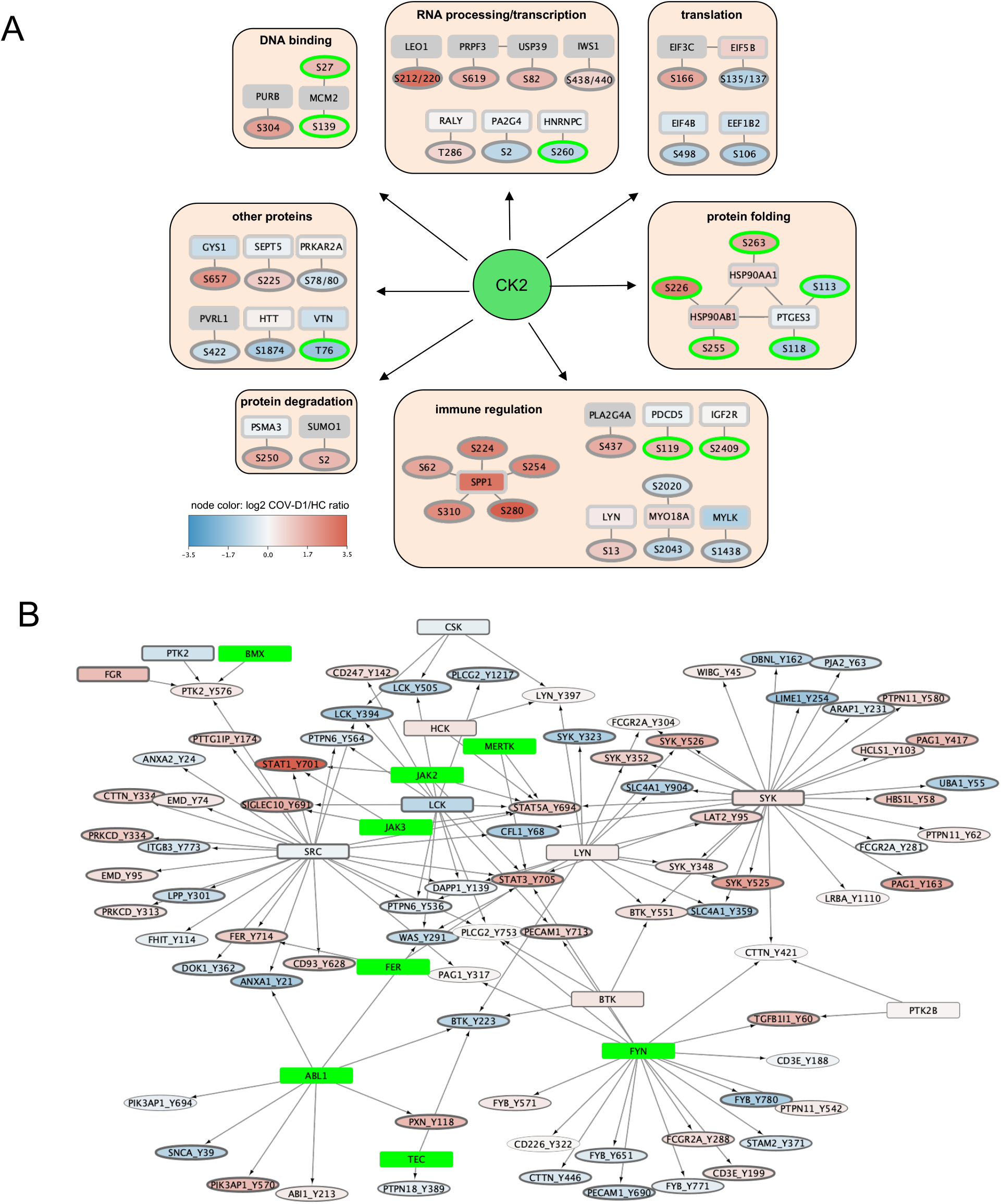
The protein kinase-substrate network underlying COVID-19. The protein kinase-substrate network underlying COVID-19 (**A**) The CK2-substrate network. Known or predicted CK2 substrate phosphosites were extracted from the significantly regulated phosphosites (ANOVA FDR < 10% between COV-D1, ICU and HC). Phosphosites with a green border are known CK2 substrates based on PhosphositePlus. The phosphorylation nodes are colored based on the IMAC data, whereas the protein nodes are based on the proteome data. Protein nodes without observation/quantification are colored grey. (B) A TK-substrate network for COVID-19 based on quantitative proteome and phosphoproteome data. The kinase nodes were coloured by the log_2_ COV-D1/HC fold change of the full proteome protein intensities. Kinase proteins which were observed by mass spectrometry but without sufficient data were colored green (proteins with n ≥ 3 from each sample group were considered sufficient for statistical analysis). The thickness of the node border correspond to p-values (thicker for smaller p-values). The kinase-substrate relationship data were retrieved from PhosphoSitePlus database (version November 2020).

Because of the central role of Tyr phosphorylation in proximal signaling by immune receptors and cytokines, we next focused our analysis on the TK-substrate network. Of the 394 quantifiable pTyr sites identified, 95 showed significant changes between COV and HC (Fig. S6A). The majority of the significantly regulated Tyr phosphorylated proteins/pTyr sites belong to a large TK-substrate signaling network (Fig. 4B). The tyrosine kinases LYN, SYK, LCK, SRC, FYN and ABL1 form the major nodes whereas BTK and JAK2/3 form the minor nodes in the network to connect the diverse array of phosphorylated substrates. Except for ABL1, all the TKs in the network were significantly more active in the COV group based on the phosphoproteome datasets or KSEA prediction. This extensive TK-substrate network suggests that SARS-CoV-2 promotes immune signaling.

### Excessive JAK-STAT signaling associated with the cytokine storm

The cytokine release syndrome (CRS) or cytokine storm is a major cause of acute lung damage associated with patient mortality^38-43^. Because the cytokine storm is also common in sepsis caused by other pathogens ^42,44^, we determined the mRNA levels of key proinflammatory cytokines and chemokines in the PBMCs of the COV (D1) and ICU cohorts in comparison to the heathy controls. Except for IL-12, IFN-α, IFN-β, and IL-4, all the cytokines/chemokines examined were significantly overexpressed in COV (Fig. 5A). This is in contrast to the ICU cohort where only a few were significantly upregulated and by a much smaller degree. SARS-CoV-2 thus elicited a much stronger cytokine storm in the COVID-19^+^ than the COVID-19^-^ ICU patients who had pneumonia and suspected sepsis (Table S1). In agreement with published data ^3,39,45,46^, the most highly expressed cytokines included TNF-α, IL-6, IFN-γ, IL-2 and IL-15. Curiously, we found the IL-1β level fairly low in comparison, consistent with a previous report^47^. Besides cytokines, the chemokines MCP-1/CCL2 (which recruits monocytes and/or macrophages) and IL-8/CXCL8 (a classic neutrophil chemoattractant), the macrophage inflammatory protein MIP-1b and the macrophage colony-stimulating factor (M-CSF) were all significantly upregulated. These data agree with the important role for neutrophils, monocytes and macrophages in the pathology of COVID-19^35,48^. In support of this assertion, iNOS, which is frequently expressed by neutrophils, macrophages and dendritic cells to produce NO, was significantly elevated. Inflammatory cytokines, including TNF-α and IFN-γ, may also induce NO production through the JAK-STAT1 axis^39^.

**Figure 5.**
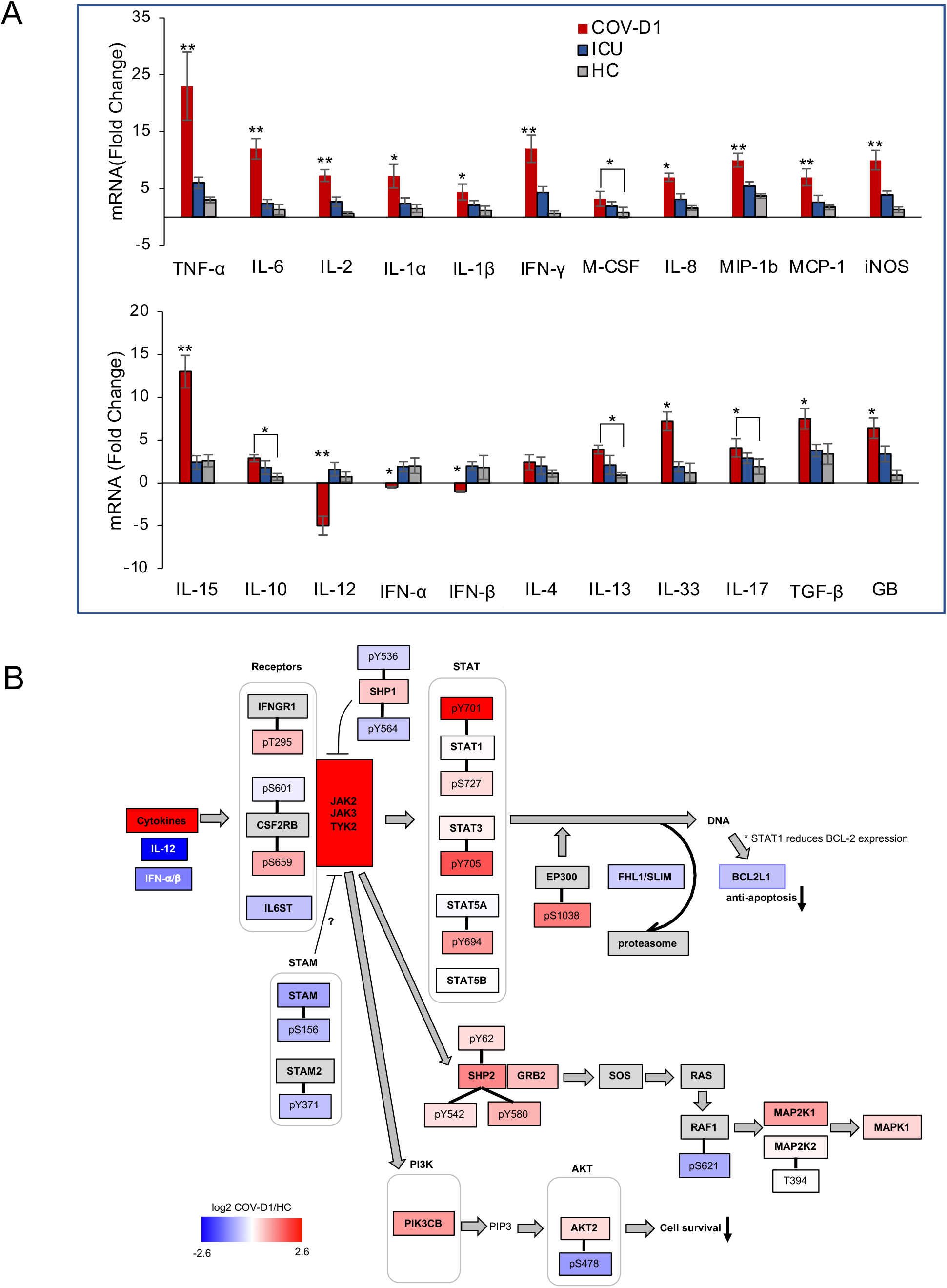
The cytokine storm and elevated JAK-STAK signaling in COVID-19 patients. (**A**) Differential expression of cytokines and chemokines in the COV-D1, ICU and HC PMBCs. Data shown were normalized to β-actin. Asterisks indicate a significant difference between COV-D1 and ICU or HC unless otherwise indicated. *, p<0.05, **, p<0.002, calculated based on One-Way ANOVA (n=5, 4 technical repeats). (**B**) The cytokine-JAK-STAT signaling network underlying COVID-19. The proteins and phosphosites are colored according to the COV-D1/HC ratio (red: upregulation, blue: downregulation). The nodes without sufficient data to calculate the ratio are colored grey.

It is intriguing that IL-12, which was slightly upregulated in ICU, was significantly down-regulated COV. In contrast, IL-10 expression, which may antagonize IL-12-mediated protection against acute virus infection^49^, is significantly increased. IL-12, produced by dendritic cells, macrophages, and lymphoblastoid cells, is required for the differentiation of Th1 cells and for the activation of NK cells, both of which have been found defective in COVID-19^46^. In contrast, the Th2 cytokines IL-4 and IL-10 were moderately increased, suggesting the CD4+ T cell lineage is skewed towards Th2 in COVID-19^46^. Besides IL-12, we found the transcripts of the type I interferons, IFN-α and INF-β, significantly reduced, suggesting impaired antiviral immunity. As discussed earlier, the suppression of IL-12 and IFN-α*/*β expression may be mediated in part by CK2-induced phosphorylation and inhibition of OPN/SPP1. Furthermore, the increased NO production via iNOS may inhibit IL-12 production by macrophages and dendritic cells ^50^. Consistent with reduced IFN-α/β signaling, we found the expression of the downstream targets ISG15 and EIF2A significantly reduced in the COV samples (Table S4).

Signal transduction by cytokines is dependent on the JAK-STAT pathway. As shown in Fig. 3C, JAK2/3 and TYK2 were the most highly activated kinases in the COV PBMCs. Network analysis indicates that the JAK2/3 activation is reinforced by decreased phosphorylation/activation of SHP1 and STAM, which are negative regulators of cytokine signaling. Activation of the Janus kinases led to significant activation of STAT1, STAT3 and STAT5A. It is interesting to note that STAT1 was the most activated STAT despite numerous reports suggesting that the IL-6-STAT3 axis plays a pivotal role in the cytokine storm^43,46^. STAT1 activation may underlie the significant changes in expression for proteins involved in regulating inflammation, complement, exocytosis, metabolism, glycosylation, cardiac myocyte survival and T cell signaling (Fig. S9). Collectively, the JAK-STAT pathway may signal to reduce the transcriptional activity of EP300 (i.e., increased S1038 phosphorylation) and the expression of FHL1/SLIM, a protein implicated in protein turnover and cardiomyopathy^51,52^. JAK2/3 may also regulate cell proliferation through the SHP2-GRB2-SOS-RAS-RAF-MEK1/2 pathway and survival through the PI3K-AKT2 pathway. The reduced phosphorylation/activation of AKT2 suggests reduced cell survival, which together with reduced BCL2L1 expression, may contribute to increased apoptosis and lymphopenia in COVID-19^39,46^.

### Elevated TCR signaling in the COVID-19 PBMCs

Immune signaling is critically dependent on immunoreceptor Tyr-based regulatory motifs (ITRMs)^53^. The phosphorylation profile of the ITRMs, therefore, provides a picture of the immune signaling landscape. Our MS analysis yielded phosphorylation data for numerous ITRMs (Fig. 6A; Table S9). Together with the quantitative expression data for many ITRM containing proteins (Fig. 6B), we are at a unique position to gauge the activity of different immune signaling pathways. Of note, five of the six pairs of immunoreceptor Tyr-based activation motifs (ITAMs) on CD3ζ, CD3δ, CD3ε and CD3γ showed increased phosphorylation in COV (Fig. 6A, 6C). This observation is consistent with the increased activation of LCK in the same samples (Fig. 3C). Given the pivotal role of the CD3 molecules in TCR signaling, this data indicates that TCR signaling is elevated in the COVID-19 patients. In accordance with this assertion, NFAM1, an activator of the calcineurin/NFAT-signaling pathway, was phosphorylated more robustly in the patient PBMCs. Collectively, these data indicate that the COVID-19 patients had highly active circulating T cells despite having lymphopenia.

**Fig. 6.**
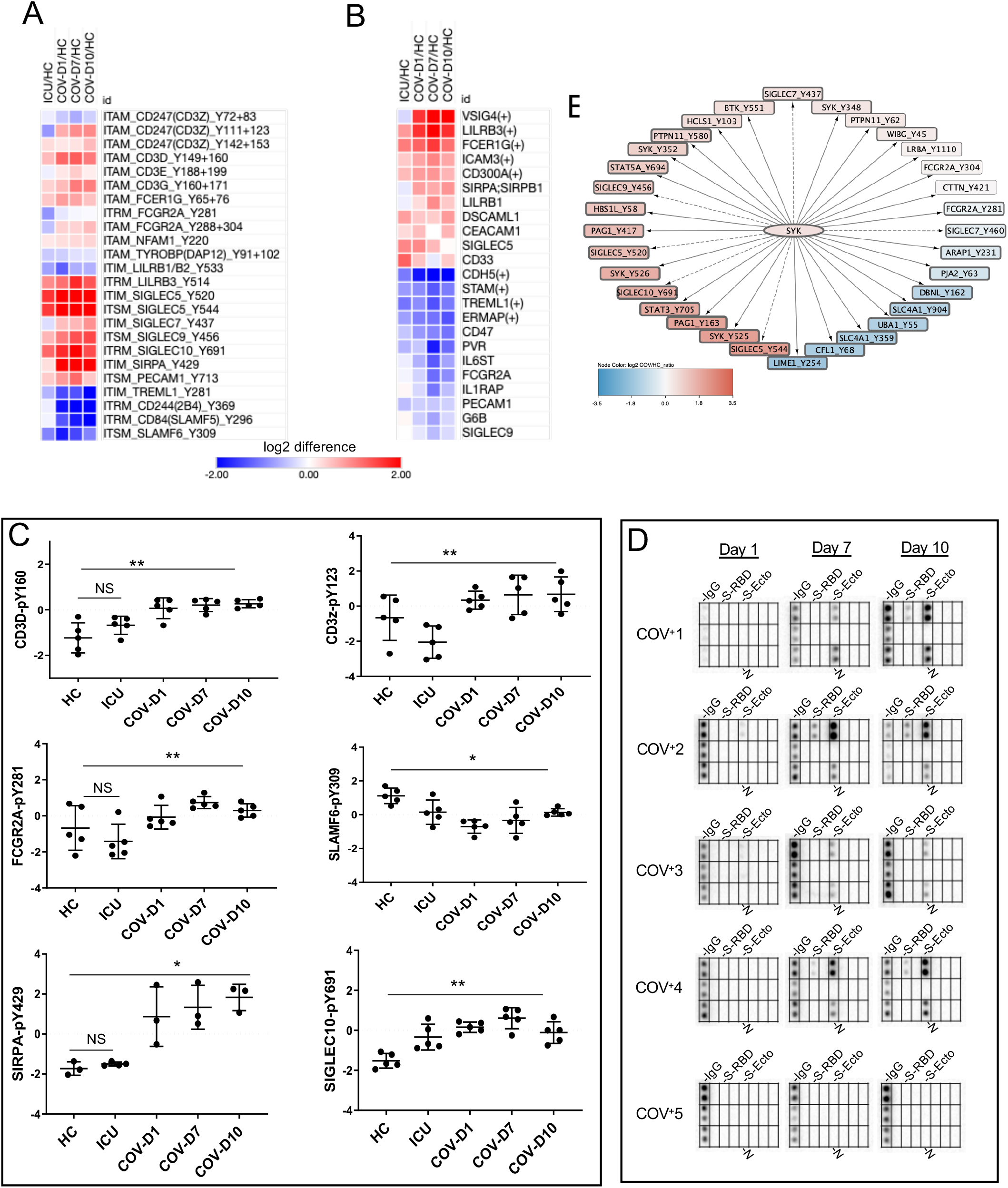
SARS-CoV-2 augments immune signaling through immunoreceptor Tyr-based regulatory motifs. (**A**) A heatmap showing the differences in ITRM Tyr phosphorylation in patient PBMCs compared to the heathy controls. (**B**) A heatmap of the ITRM-containing proteins with significant changes in expression. (**C**) Representative examples showing the dynamic changes in phosphorylation of the ITRMs. p-values calculated by One-way ANOVA (n=3-5), *, p<0.05, **, p<0.002. (**D**) Antibody response determined by a SARS-CoV-2 antigen array. (**E**) SYK may phosphorylates multiple proteins and nucleate an immune signaling hub. The SIGLEC pTyr sites with dashed arrows are potential SYK substrates.

### Compromised NK cell signaling

Contrary to elevated T cell signaling, we found that signaling by NK cells, which may kill virus-infected cells, were significantly compromised in COV. The activity of NK cells is regulated by the SLAM family receptors that contain ITSM sequences^54^. We found that the Tyr phosphorylation of several SLAM members, including SLAM6, CD84 and CD244 (2B4), significantly reduced. Moreover, phosphorylation of the ITAM in TYROBP/DAP-12, an adaptor for the activating NK cell receptor CD94/NKG2C, was down-regulated in both the COV and ICU cohorts. Moreover, both CD300A/MAIR-1, an inhibitory receptor in NK cells, and CEACAM1, a coinhibitory receptor in T cells, NK cells and neutrophils, were increased in expression (Fig. 6B). In contrast, the expression of PVR, which mediates NK cell adhesion and triggers NK cell effector functions, was significantly reduced (Fig. 6B). Collectively, these data suggest that NK cell receptor-mediated signaling and effector function were compromised by SARS-CoV-2. The defective NK cell signaling is consistent with reduced IL-12 expression as IL-12 plays a critical role in NK cell activation.

### Antibody response and Fc receptor-mediated phagocytosis

Phagocytosis is an important defense mechanism against pathogen infection. The elimination of infected cells or pathogens can be facilitated by Fc receptor-dependent phagocytosis. Consistent with up-regulation of the FcR pathway (Fig. 2), signaling through the Fcε receptor FCER1G ITAM was seen elevated in both patient groups whereas the Fcγ receptor FCGR2A ITAM phosphorylation was significantly upregulated in the COV but not the ICU group (Fig. 6A & 6C). To find out if this was caused by virus specific IgG, we determined the antibody response to SARS-CoV-2 using an antigen array containing recombinant viral proteins (Fig. S10). Probing the protein antigen array by the COVID-19 patient plasma yielded the antibody response profiles (Fig. 6D). Four of the five COVID-19 patients showed robust anti-Spike and anti-nucleocapsid antibodies in the plasma collected on day 7 and day 10 of ICU admission. No antibody was detected for the fifth patient (COV+5) using the same array, but antibody response to the Spike ecto-domain was detectable when the antigen concentration was increased by 10-fold (Fig. S11). Intriguingly, this patient was the only one who recovered from the SARS-CoV-2 infection while the remaining four succumbed to the disease. This suggests that robust antibody response is not an indicator of potent immunity against the virus, as have been shown in previous studies^55^.

A possible reason behind this phenomenon is compromised phagocytosis due to co-inhibitory signaling mediated by the immunoreceptor based inhibitory or switch motifs (ITIMs/ITSMs). While FcR signaling positively regulates phagocytosis by phagocytes, it can be countered by inhibitory receptors that bear the ITIM/ITSM motifs. A number of ITIM-containing receptors were found indeed activated, presumably in neutrophils or macrophages. Of note, phosphorylation of Y429 in the inhibitory receptor SIRPα, was significantly up-regulated (Fig. 6A & 6C). Coincidently, the expression of SIRPα was significantly increased in COV, but not ICU (Fig. 6B). PECAM1, another inhibitory receptor abundantly expressed on macrophages, was also significantly activated (Fig. 6A). This suggests SIRPα and PECAM1 may work together to impede phagocytosis by neutrophils and macrophages^56^. Intriguingly, we found that TREML1, a member of the TREM (triggering receptor expressed on myeloid cells) family, was down-regulated in both expression and ITIM phosphorylation. Furthermore, ITAM phosphorylation in TYROBP/DAP-12, an adaptor in TREM1 and TREM2 signaling^57^, was significantly reduced. Collectively, these data suggest that the Fc receptor-mediated phagocytosis via macrophages or neutrophils is compromised by robust inhibitory signaling in the COV group.

SIGLECs are a family of receptors that play an important role in immune self-tolerance and host defense^58^. Different SIGLECS are expressed in different myeloid and lymphatic cells and plays a negative role in regulating the effector function of these cells. We found that ITIM/ITSM phosphorylation in SIGLEC5, SIGLEC7, SIGLEC9 and SIGLEC10 all increased in the COV cohort. Because the SIGLECs are found in neutrophils, monocytes, NK cells and B cells, this suggests immune tolerance in the COVID-19 patients may be mediated by enhanced sialic acid signaling in these cells. Furthermore, increased inhibitory signaling via the SIGLECs suggests compromised myeloid cell function. Finally, it is worth noting that SYK may be a potential target for intervention due to its ability to phosphorylate numerous substrates in immune cells^59^, including the SIGLECs, PAG1-a negative regulator of T cell signaling, PTPN11/SHP-2-negative regulator of immune signaling, and STAT3/STAT5, key players in cytokine signaling (Fig. 6E).

## DICUSSION

Our deep and quantitative MS analysis uncovered the largest proteome and phosphoproteome for the COVID-19 patient peripheral blood derived mononuclear cells (PBMCs) to date. This allowed for comparison of SARS-CoV-2^+^ ICU patients with age- and sex-matched SARS-CoV-2^-^ ICU patients and heathy individuals at the proteome, phosphoproteome, network and pathway levels. Besides significant differences in the phagocytosis, complement/coagulation, platelet activation, metabolism, antigen presentation, and cytokine signaling pathways, the COVID-19 PMBCs are characterized with a highly active kinome and extensive kinase-substrate networks. Our work shows that severe COVID-19 is marked with a highly active adaptive immune response, compromised innate immune response, and imbalance between antiviral and proinflammatory responses ^46^.

### T cell and B cell activation despite of lymphopenia

Despite a general reduction in lymphocyte count, robust T cell and B cell subsets have been reported in a proportion of patients^5,60,61^. We showed that the critically ill COVID-19 patients had highly activated T cell and B cell signaling that are regulated by the tyrosine kinases LCK, LYN and SYK. The T cell activation was manifested in increased ITAM phosphorylation in the CD3 proteins. At the same time, negative regulators of TCR signaling, including DOK, PAG, and CSK were inhibited in the COVID-19 PBMCs. It is likely that the TCR was activated be pMHC-I as MHC-II mediated antigen presentation was deficient in COVID-19 (Fig. 2) ^62^. Because DCs are a major source of IL-12 production, the reduced IL-12 expression observed in the COV cohort may reflect reduced dendritic cell numbers in the critically ill COVID-19 patients. Similarly, the activated TCR and BCR signaling is consistent with reports that the magnitude and functional breadth of virus-specific CD4 T cell and antibody responses are consistently higher among hospitalized patients^63^.

### Enhanced inhibitory signaling and impaired innate immunity

We have shown that the impairment in innate antiviral immunity involves at least three mechanisms. First, SARS-CoV-2 compromises phagocytosis by promoting inhibitory signaling in phagocytes. Activation of the ITIM/ITSM-containing inhibitory receptors SIRPα and PECAM1 may contribute to reduced phagocytosis whereas activation of the SIGLECs may block phagocytosis by a wide range of professional phagocytic cells, including neutrophils, monocytes, dendritic cells and macrophages. By countering FcR-mediated phagocytosis, the sialic acid-SIGLEC axis may be exploited by SARS-CoV-2 to promote immune tolerance^64^ or even contribute to antibody-dependent enhancement ^65^. Potential sialic acid or galactose binding domains have been described in the Spike protein by docking analysis^66^. The broad-scale impairment of phagocytosis mediated by the ITIM/ITSM-containing inhibitory receptors likely explains why patients who developed neutralizing antibodies earlier in infection had a higher rate of the disease and worse outcome than those who did not ^67,68^. Second, SARS-CoV-2 interferes with NK cell signaling and effector function. A number of studies have reported the association of reduced NK cell number and disease severity. Moreover, ex vivo NK cells from the peripheral blood of COVID-19 patients show impaired cytotoxicity^69^. We have showed that signaling through the SLAM family receptors, which are important regulators of NK cell activity, is significantly inhibited. Besides reduced FYN activation (Fig. 3A), which is responsible for SLAM receptor phosphorylation, IL-12 deficiency may contribute significantly to reduced NK cell activity. Third, SARS-CoV-2 inhibits the production of the type I interferons IFN-α/β ^70^. Because plasmacytoid dendritic cells (pDCs) are the major source of IFN-α*/*β and IL-12, reduced expression of these cytokines is consistent with previous reports showing reduced pDC function and defects in MHC-II-dependent antigen presentation^62^.

### Targets for potential immunomodulatory therapy

In addition to providing information on the regulatory mechanisms of adaptive and innate immune response, our work has identified a number of therapeutic targets, including CK2, SYK, JAK3, TYK2, DNA-PK and IL-2. CK2 is likely a master regulator of immune response in COVID-19. CK2 may directly regulate viral RNA sensing and antiviral defense via the CK2-RIG1-TKB1-IRF3-IFN-α*/*β pathway. CK2 knockdown or inhibition has been shown to activate RIG1, TKB1 and IFN-α/β response^71,72^. Our work suggests that CK2 may also regulate type I interferon response via the CK2-OPN-IFNα/β axis and inactivate NK cell effector function via CK2-OPN-IL-12. Furthermore, CK2 activation may impact the JAK-STAT pathway by phosphorylating JAK2^73^. Therefore, by inhibiting CK2, it is possible to rejuvenate antiviral immunity and at the same time, reduce the damaging effect of the cytokine storm. In that vein, it is worth noting the CK2 inhibitor silmitasertib suppressed SARS-CoV-2 infection in vitro in cell model ^8^.

SYK is another attractive target emerging from our work ^74^. SYK activation in COVID-19 may be a double-edged sword^75^. On the one hand, it is required for the B-cell receptor and FcR-mediated signaling pathways, and on the other hand, it is involved in promoting inhibitory pathways in innate immune cells^59^, thereby compromising FcR-mediated phagocytosis to eliminate the virus or virus-infected cells. In the latter capacity, it is possible that SYK may even contribute to ADE. Indeed, SYK has a crucial role in autoimmune diseases and haematological malignancies^76^. SYK inhibitors may thus alleviate the autoimmune response associated with COVID-19^29^.

Lastly, we propose that IL-12 supplementary therapy alone or in combination with proinflammatory cytokine blockade may be effective strategies to treat critically ill patients. The wide ranges of cytokines that are significantly overexpressed, which may signal to different STAT proteins to affect a wide range of cellular functions, may not be overcome by targeting a single cytokine-receptor pair. For example, there are at least 9 other cytokines of the IL-6 family that can activate STAT3 besides IL-6 itself ^41^. This may explain why tocilizumab, an inhibitor of the IL-6 receptor, did not show significant benefits on disease progression or survival of hospitalized patients with COVID-19 pneumonia in clinical trials^77,78^. Indeed, anticytokine therapies in severe COVID-19 should be informed by detailed inflammatory profiling^79^ and be applied according to the underlying molecular mechanisms. Our work suggests that IL-12 should be included in future cytokine-based therapies. Furthermore, JAK3, JAK2 and TYK2, which are highly activated in COVID-19, may be explored. Because these kinases transduce signals downstream of the cytokines, the corresponding inhibitor may help alleviate ARDS associated with CRS.

The pathophysiology of SARS-CoV-2-induced ARDS has similarities to that of severe community-acquired pneumonia and sepsis caused by other viruses or bacteria ^40^, as is the case between the COV and ICU cohorts employed in the current study. The rich proteomics and phosphoproteomics datasets obtained herein may inform future investigations into the mechanisms underpinning pneumonia and sepsis associated with virus or bacterium infection and aid in the development of targeted immunomodulatory therapies for the treatment of these conditions^39^. Furthermore, quantitative MS analysis of the peripheral blood, a readily available biospecimen, may be used to evaluate individual responses to COVID-19 therapies or vaccines.

## Supporting information

checklist

## Data Availability

The mass spectrometry data are available from the PRIDE databse accession PXD024087.

## ACKNOWLEDGEMENT

The proteins used in the proteome array were supplied by the Toronto Open Access COVID-19 Protein Manufacturing Center (comprising BioZone and the Structural Genomics Consortium (SGC)) under an Open Science Trust Agreement (http://www.thesgc.org/click-trust). These included the SARS-CoV-2 Spike-ectodomain, nucleocapsid-dimerization, nucleocapsid-RNA binding domain, NSP3-unique, NSP3-ADRP, NSP3-NAB, NSP3-PLPro, NSP4-CTD, NSP5, NSP7, NSP8, NSP9, NSP10, NSP16. The Center received funding from the Toronto COVID-19 Action Fund. We acknowledge funding from the Ontario Research Fund-COVID-19 Rapid Research Fund, Western University (Research), the Departments of Medicine and Pediatrics at Western University, the Lawson Health Research Institute (https://www.lawsonresearch.ca/), the London Health Sciences Foundation (https://lhsf.ca/), and the AMOSO Innovation Fund. SE was supported by a Post-Doctoral Fellowship from the National Science and Engineering Council of Canada. SSCL holds a Canada Research Chair in Molecular and Epigenetic Basis of Cancer.

## MAIN FIGURE TITLES AND LEGENDS

Each spot corresponds to a protein within the specified pathway based on GO annotation. The y-axis shows the log2 value of the fold change in protein expression for the group (relative to the mean of the whole proteome). The p values were calculated based on ANOVA test, *p<0.05, **p<0.002, NS, not significant.

## SUPPLEMENTAL FIGURE TITLES AND LEGENDS

**Fig. S1. An overview of sample processing for LC/MS-MS analysis**.

PMBCs were isolated from patients or healthy control blood samples. Another PBMC sample was treated with pervanadate to be used as pTyr booster in the 11-plex TMT experiments. Protein materials were isolated and digested by LysC and trypsin. The samples were labelled with 3 sets of 11-plex TMT labelling reagents. The 11 peptide samples for each isobaric set were combined and incubated with SH2 superbinder beads. The bound peptides were enriched with tyrosine-phosphorylated peptides. The flow-though materials were used for analysis of the full proteome and the phosphoproteome (mainly for pSer and pThr peptides). The Ti4+-IMAC resin was used for the phosphopeptide enrichment. Proteome peptides and enriched phosphopeptides were fractionated using a high-pH fractionation kit prior to LC-MS/MS analysis.

**Fig. S2. Proteins differentially expressed in COVID-19+ ICU patients compared to healthy controls**.

A Volcano plot comparing the identified PBMC proteome between the COVID-19 patients (day 1 samples) and healthy controls. 521 proteins (marked red) were found significantly different between the two sample groups (FDR 5%).

**Fig. S3. PPI network clustering by Metascape on the differentially expressed proteins in the COVID-19 PBMCs**.

(A) A summary of enriched networks and the associated top 3 functional annotation terms. MCODE_1 to 4 correspond to the depicted PPI network 1-4 below. (B-E) PPI networks significantly enriched in the COVID-19 PMBCs. A color gradient from cyan to orange was used to indicate decreased to increased expression between the COV-D1 and HC groups with the dot size indicating Log2 of the difference (from −2.7 to 2.5).

**Fig. S4. Gene Ontology (GO) enrichment analysis of proteins differentially expressed between patients and healthy controls**.

(A) Enriched processes or functions for the Cluster 1 proteins (in Fig. 1A). (B) Enriched processes or functions for the Cluster 5 proteins (in Fig. 1A).

**Fig. S5. Distinguishing features of the COVID-19 PBMC phosphoproteome compared to that of the heathy controls or ICU patients**.

Biological processes significantly enriched between (A) COVID-Day1 and HC, and, (B) COVID Day1 and ICU based on PHOTON analysis. The enrichment score and FDR (Benjamini-Hochberg) q-value from Fisher’s exact test are shown on the y-axis.

**Fig. S6. The Tyr phosphoproteome signature of COVID-19**.

(A) Heatmap showing clustering of significantly regulated pTyr sites. The 94 significantly regulated pTyr sites were identified based on the ANOVA test for the three sample groups with 10% FDR cut-off, and were grouped by hierarchical clustering. (B) Enriched GO categories for all proteins displaying significant pTyr changes between groups.

**Fig. S7. Kinase protein abundance in PBMC samples from patients relative to healthy controls**.

The scale is in log2 ratio between the sample groups. (A) All tyrosine kinases. (B) All kinases with ANOVA FDR < 10% between the three groups (COV-D1, ICU, HC).

**Fig. S8. OPN/SPP1 phosphorylation is significantly upregulated in the COVID-19 PBMCs**.

(A) A volcano plot comparing the identified pSer/pThr sites between the COV-D1 and HC1 samples. The OPN/SPP1 phosphorylation sites are identified in red. (B) A heatmap showing the intensities of all SPP1 phosphosites in the patient PBMCs relative to the healthy control. The significant pSer sites are identified by asterisks.

**Fig. S9. STAT1 regulated gene expression in the COV and ICU patients**.

Log2 fold change in expression relative to the HC group is shown. The heatmap is generated color gradient from blue to red, representing decreased to increased (Log2 fold change) expression. Significantly regulated proteins are identified in asterisks with brief functional annotation on the right.

**Fig. S10. SDS PAGE images of the recombinant SARS-CoV-2 proteins used in the antigen array**.

**Fig. S11. SARS-CoV-2 Proteome antigen array**.

(A) Identity and position of proteins printed in the proteome array (see also Fig. 5). (B) The plasma from the ICU (COVID-19 negative) and healthy control group showed no antibody response to any virus antigen. (C) A proteome array printed with 10 x spike Ecto domain (5 μM) was screened with the plasma samples from the COV+5 patient.

## SUPPELMENTAL TABLE TITLES

**Table S1. Subject demographics and clinical data**

**Table S2. The list of samples for TMT labelling and mass spectrometry analysis**

**Table S3. Mass spectrometer data collection parameters**

**Table S4. Identified proteins**

**Table S5. Identified phosphosites by IMAC enrichment**

**Table S6. Identified tyrosine phosphorylation sites by superbinder enrichment**

**Table S7. Identified phosphosites from protein kinases**.

ANOVA test was conducted between the three groups of COV-D1, ICU, HC.

**Table S8. Known or predicted CK2 substrates**.

Only the significantly regulated phosphosites are included (ANOVA FDR < 10% based on comparison between the three groups, COV-D1, ICU and HC).

**Table S9. Phosphorylated immunoreceptor tyrosine-based regulatory motifs**.

All pTyr sites of proteins that possess an ITRM motif are included here. The ANOVA test was conducted between the three sample groups (COV-D1, ICU, HC).

