## Supplementary Figures for "System-wide hematopoietic and immune signaling aberrations in COVID-19 revealed by deep proteome and phosphoproteome analysis"

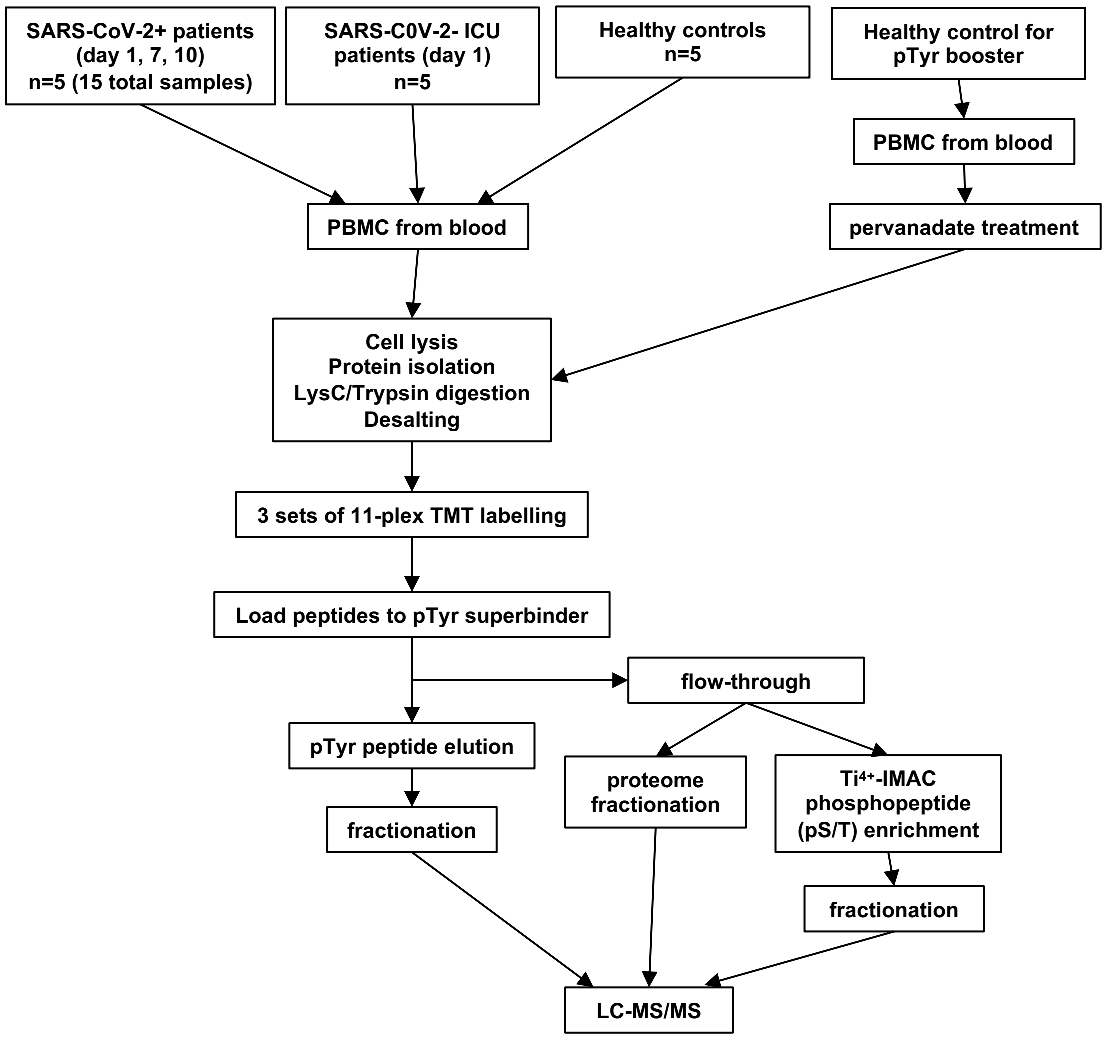


**Fig. S1. An overview of sample processing for LC/MS-MS analysis**.

PMBCs were isolated from patients or healthy control blood samples. Another PBMC sample was treated with pervanadate to be used as pTyr booster in the 11-plex TMT experiments. Protein materials were isolated and digested by LysC and trypsin. The samples were labelled with 3 sets of 11-plex TMT labelling reagents. The 11 peptide samples for each isobaric set were combined and incubated with SH2 superbinder beads. The bound peptides were enriched with tyrosine-phosphorylated peptides. The flow-though materials were used for analysis of the full proteome and the phosphoproteome (mainly for pSer and pThr peptides). The $Ti^{4+}$-IMAC resin was used for the phosphopeptide enrichment. Proteome and enriched phosphopeptides were fractionated using a high-pH fractionation kit prior to LC-MS/MS analysis.

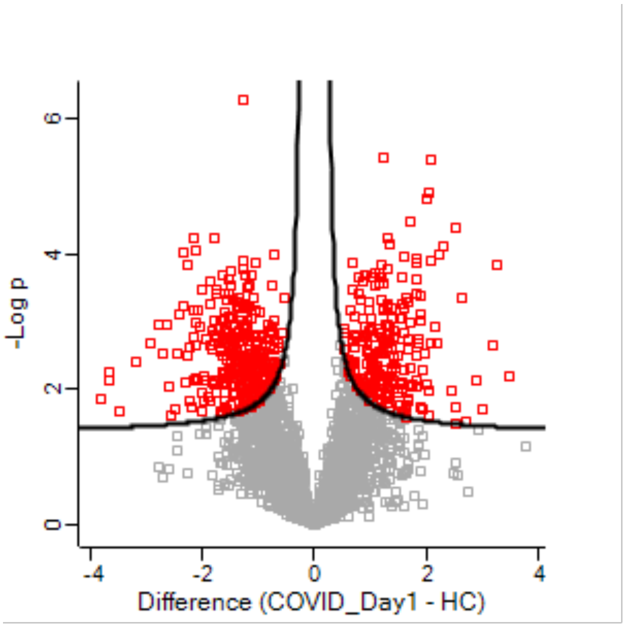


**Fig. S2**. **Proteins differentially expressed in COVID-19+ ICU patients compared to healthy controls**. A Volcano plot comparing the identified PBMC proteome between the COVID-19 patients (day 1 samples) and healthy controls. 521 proteins (marked red) were found significantly different between the two sample groups (FDR 5%).

A

| MCODE | GO | Description | Log10(P) |
|---|---|---|---|
| MCODE_1 | GO:0044772 | mitotic cell cycle phase transition | -8.7 |
| MCODE_1 | GO:0044770 | cell cycle phase transition | -8.3 |
| MCODE_1 | ko04340 | Hedgehog signaling pathway | -7.9 |
| MCODE_2 | R-HSA-8953854 | Metabolism of RNA | -10.6 |
| MCODE_2 | ko03040 | Spliceosome | -9.4 |
| MCODE_2 | hsa03040 | Spliceosome | -9.4 |
| MCODE_3 | GO:0034655 | nucleobase-containing compound catabolic process | -11.7 |
| MCODE_3 | GO:0046700 | heterocycle catabolic process | -11.3 |
| MCODE_3 | GO:0044270 | cellular nitrogen compound catabolic process | -11.3 |
| MCODE_4 | GO:0045055 | regulated exocytosis | -8.6 |
| MCODE_4 | R-HSA-2262752 | Cellular responses to stress | -6.8 |
| MCODE_4 | R-HSA-8953897 | Cellular responses to external stimuli | -6.8 |

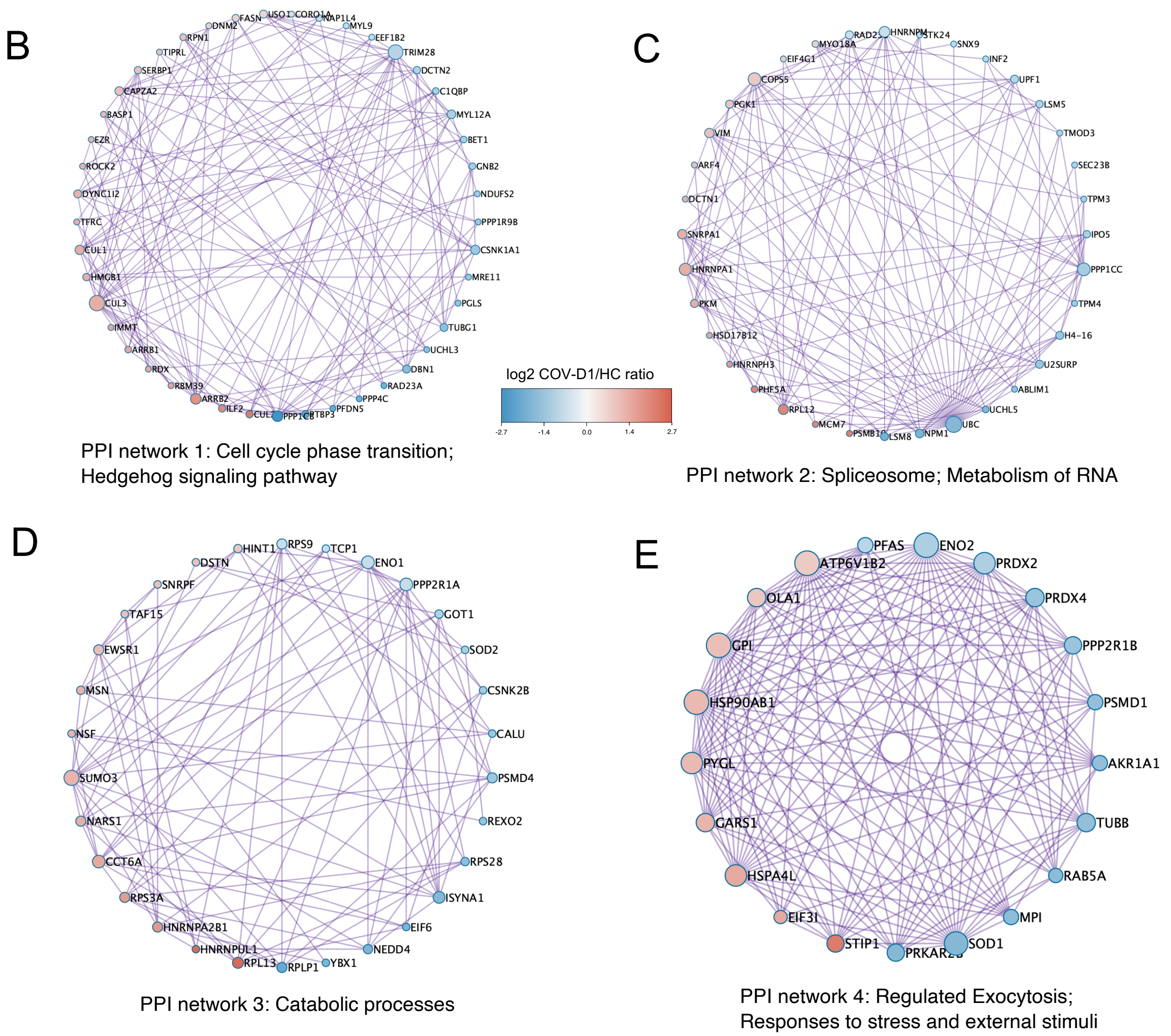


PPI network 1: Cell cycle phase transition; Hedgehog signaling pathway

PPI network 2: Spliceosome; Metabolism of RNA

PPI network 3: Catabolic processes

PPI network 4: Regulated Exocytosis; Responses to stress and external stimuli

**Fig. S3. PPI network clustering by Metascape on the differentially expressed proteins in the COVID-19 PBMCs**. (A) A summary of enriched networks and the associated top 3 functional annotation terms. MCODE_1 to 4 correspond to the depicted PPI network 1-4 below. (B-E) PPI networks significantly enriched in the COVID-19 PMBCs. A color gradient from cyan to orange was used to indicate decreased to increased expression between the COV-D1 and HC groups with the dot size indicating Log2 of the difference (from -2.7 to 2.5).

A

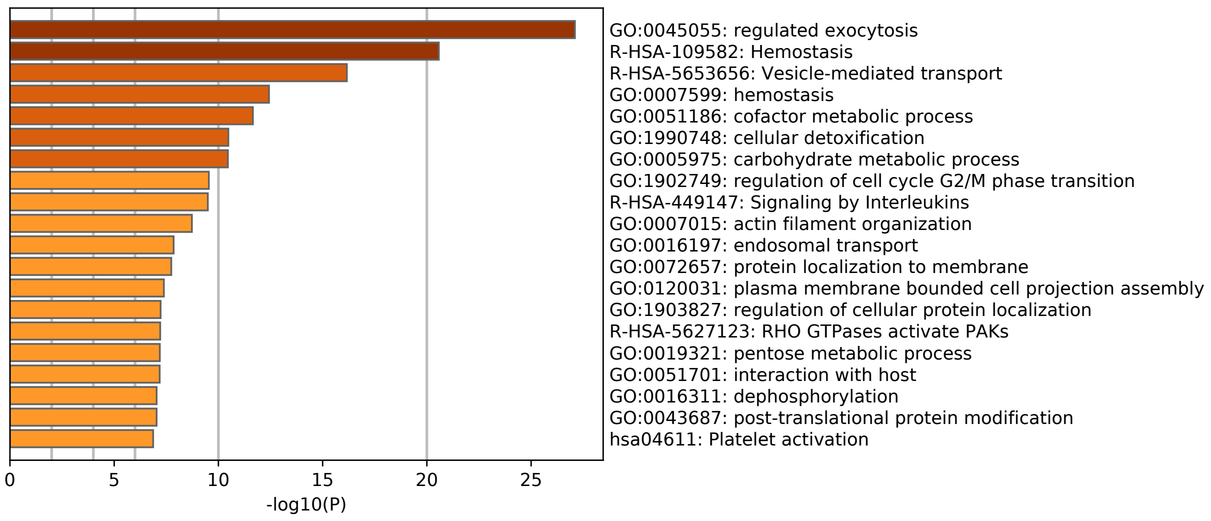


B

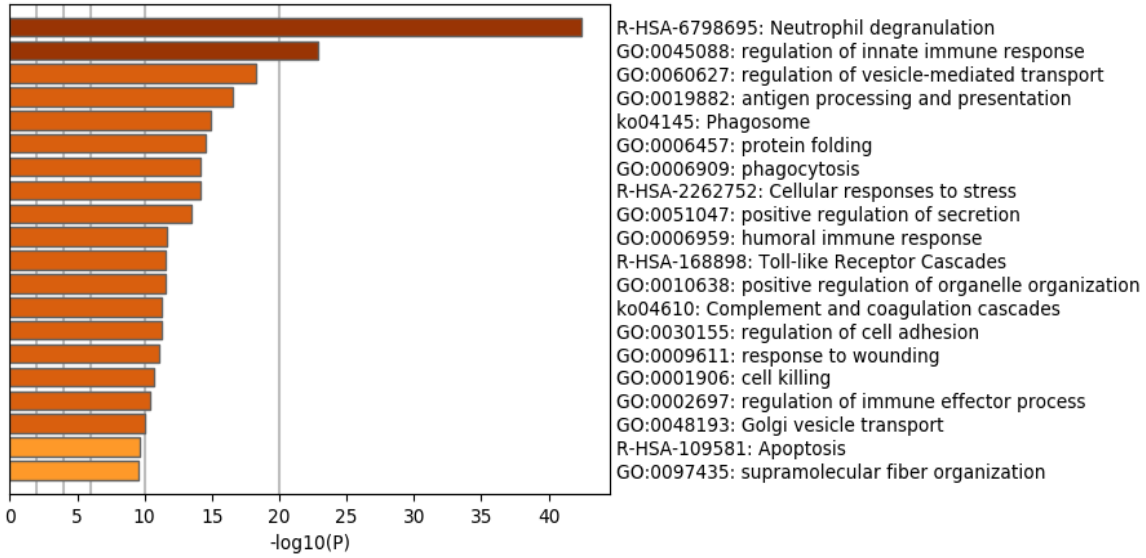


**Fig. S4. Gene Ontology (GO) enrichment analysis of proteins differentially expressed between patients and healthy controls.** (**A**) Enriched processes or functions for the Cluster 1 proteins (in Fig. 1A). (**B**) Enriched processes or functions for the Cluster 5 proteins (in Fig. 1A).

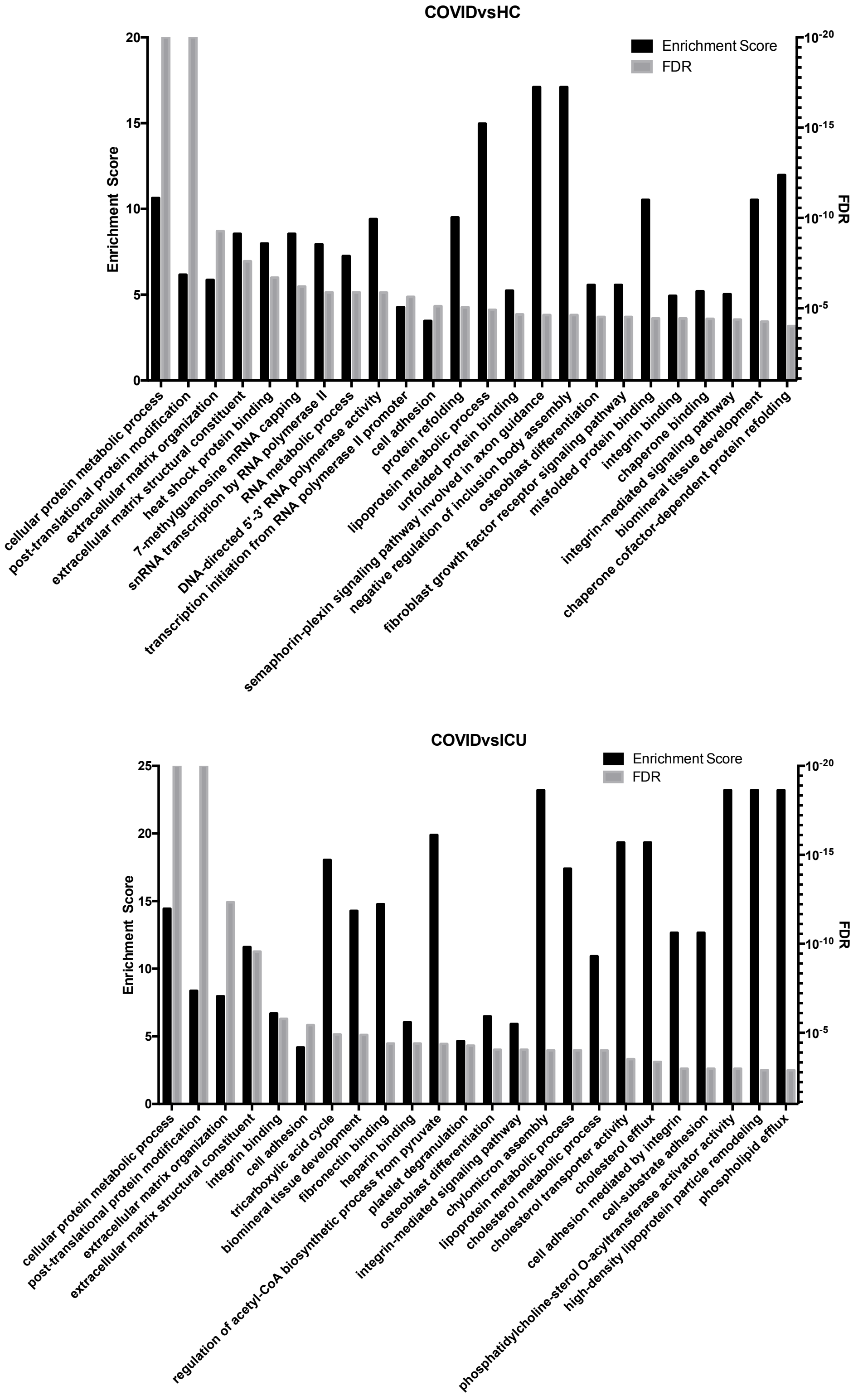


**Fig. S5. Distinguishing features of the COVID-19 PBMC phosphoproteome compared to that of the heathy controls or ICU patients**. Biological processes significantly enriched between (**A**) COVID-Day1 and HC, and, (**B**) COVID Day1 and ICU based on PHOTON analysis. The enrichment score and FDR (Benjamini-Hochberg) *q*-value from Fisher’s exact test are shown on the y-axis.

A

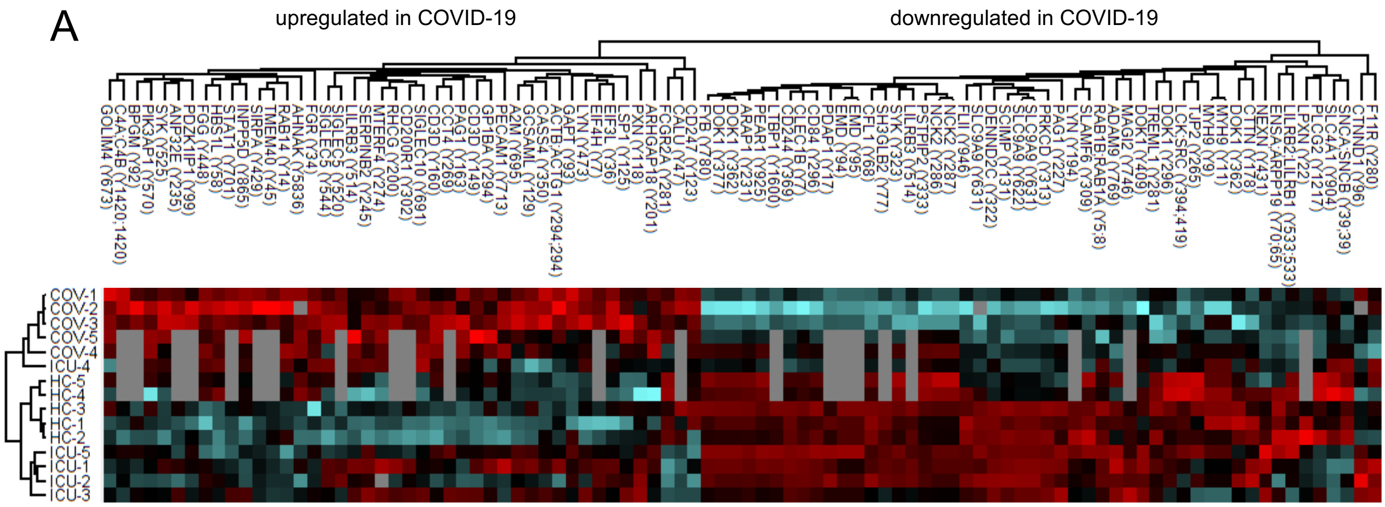


B

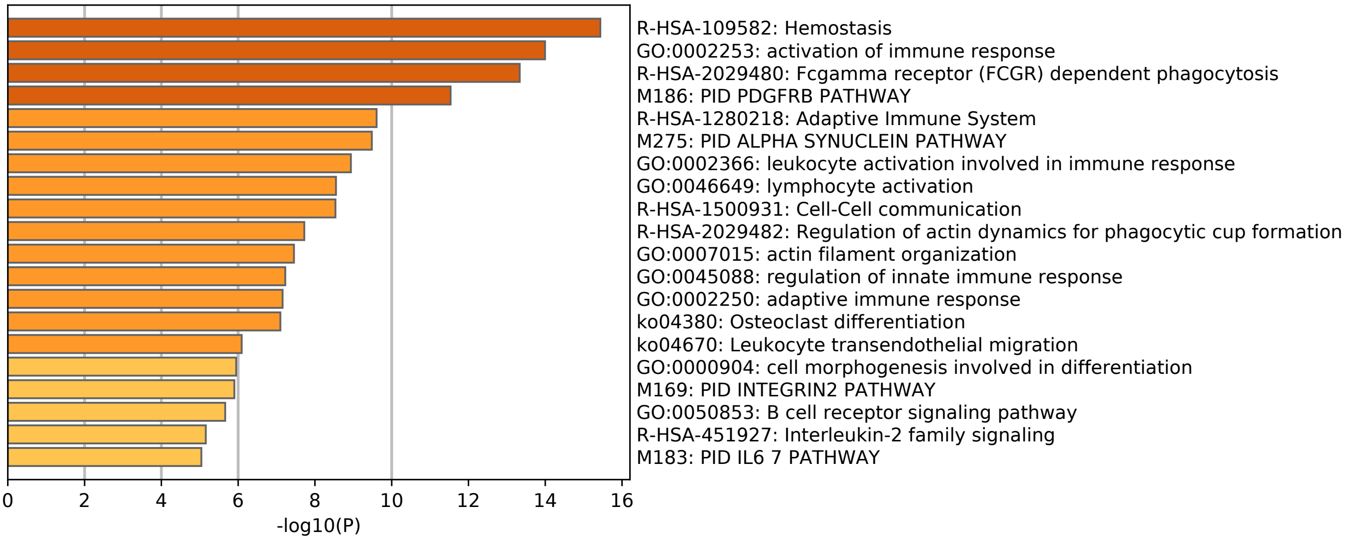


**Fig. S6. The Tyr phosphoproteome signature of COVID-19.** (A) Heatmap showing clustering of significantly regulated pTyr sites. The 94 significantly regulated pTyr sites were identified based on the ANOVA test for the three sample groups with 10% FDR cut-off, and were grouped by hierarchical clustering. (B) Enriched GO categories for all proteins displaying significant pTyr changes between groups.

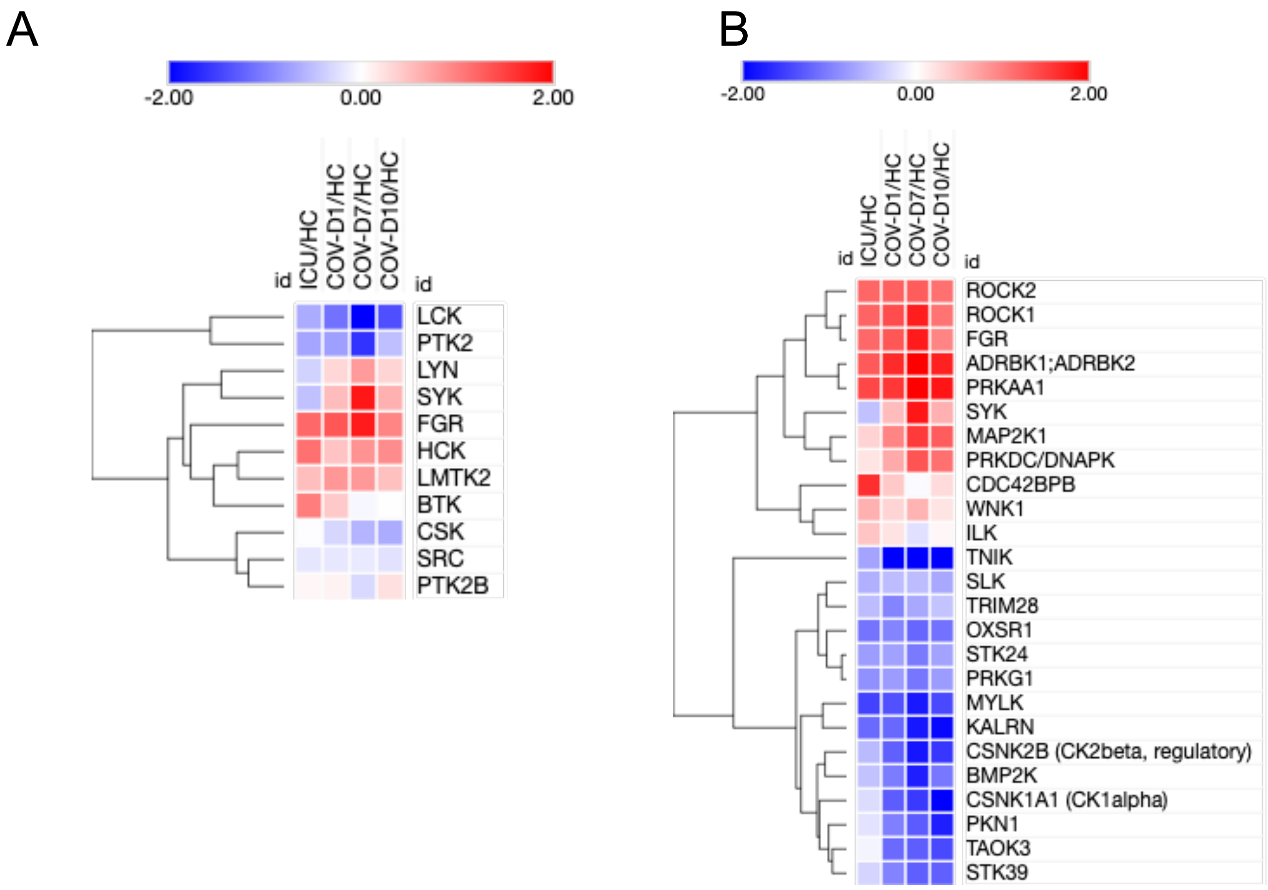


**Fig. S7. Kinase protein abundance in PBMC samples from patients relative to healthy controls**. The scale is in log2 ratio between the sample groups. (A) All tyrosine kinases. (B) All kinases with ANOVA FDR < 10% between the three groups (COV-D1, ICU, HC).

A

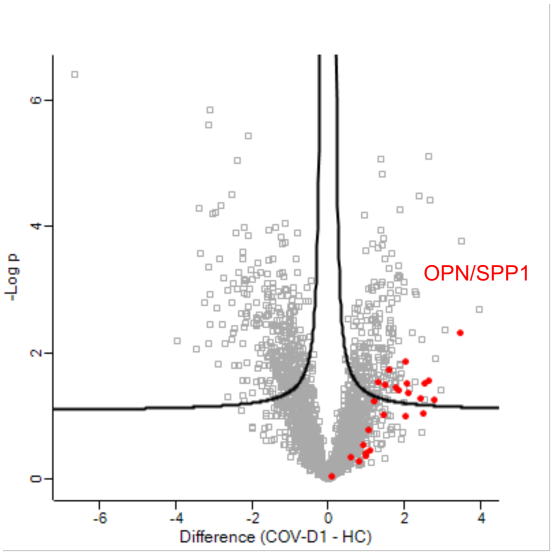


B

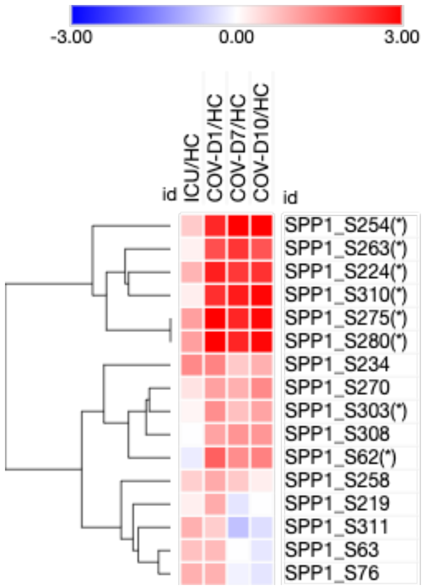


**Fig. S8. OPN/SPP1 phosphorylation is significantly upregulated in the COVID-19 PBMCs**.
(A) A volcano plot comparing the identified pSer/pThr sites between the COV-D1 and HC1 samples. The OPN/SPP1 phosphorylation sites are identified in red. (B) A heatmap showing the intensities of all SPP1 phosphosites in the patient PBMCs relative to the healthy control. The significant pSer sites are identified by asterisks.

**Fig. S9. STAT1 regulated gene expression in the COV and ICU patients**. Log2 fold change in expression relative to the HC group is shown. The heatmap is generated color gradient from blue to red, representing decreased to increased (Log2 fold change) expression. Significantly regulated proteins are identified in asterisks with brief functional annotation on the right.

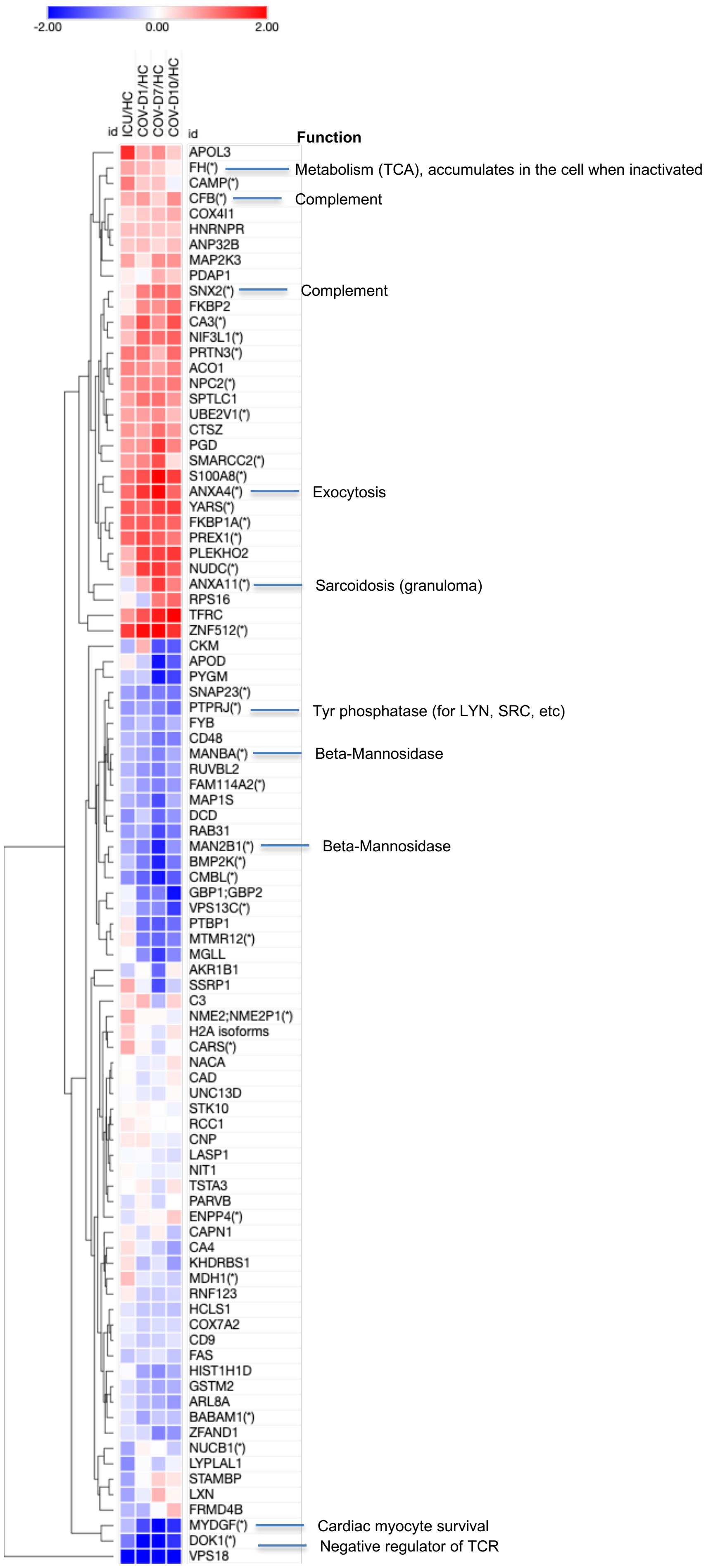

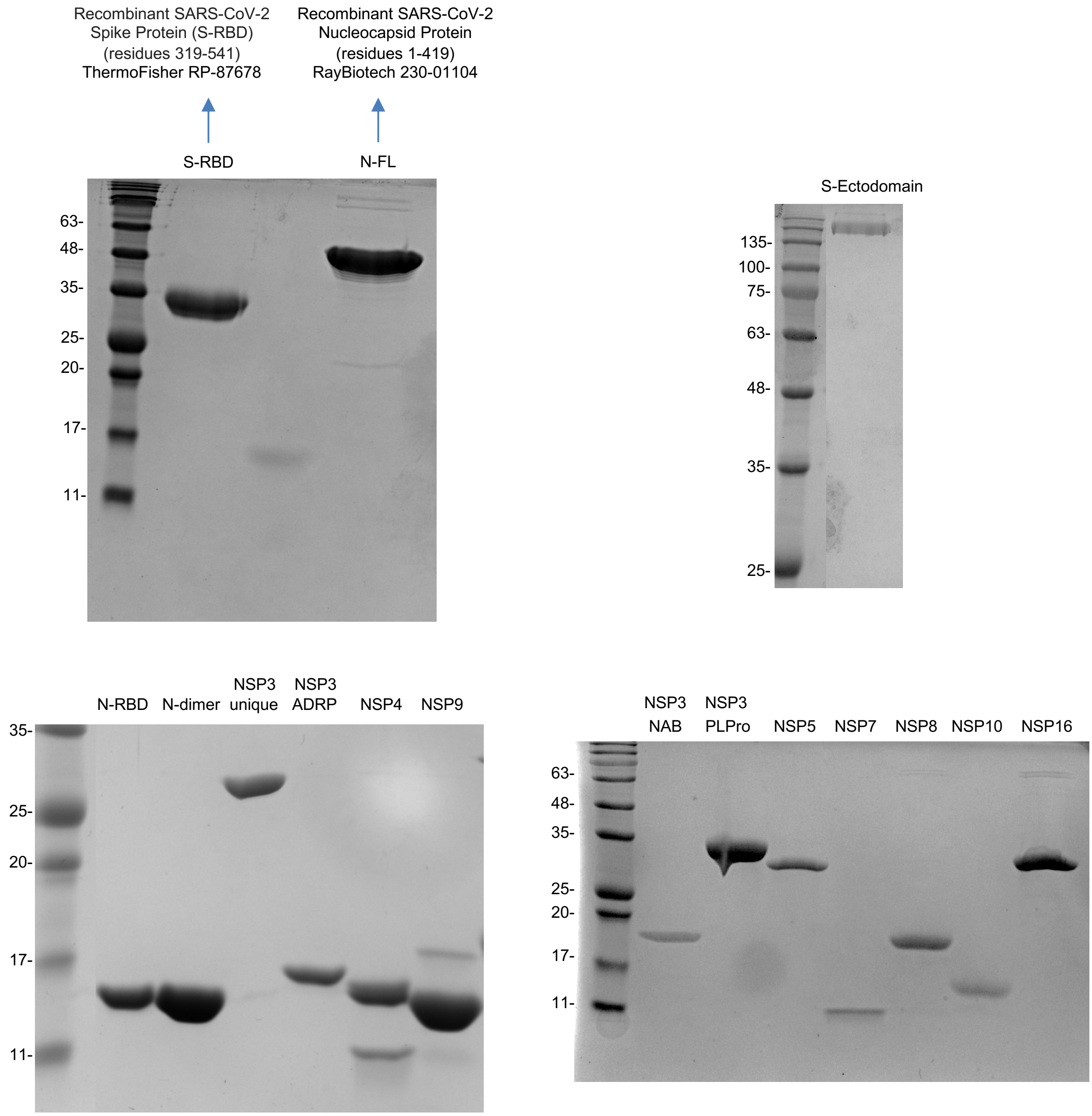


**Fig. S10. SDS PAGE images of the recombinant SARS-CoV-2 proteins used in the antigen array.**

A

| | | | | | | | |
|---|---|---|---|---|---|---|---|
| IgG | PBS | S-RBD | PBS | S-Ecto | PBS | NSP3-u | NSP3-ADRP |
| IgG | NSP3-PLPro | NSP3nab | NSP4 | NSP5 | NSP7 | NSP8 | NSP9 |
| IgG | NSP10 | NSP16 | PBS | N-FL | PBS | N-Dimer | N-RBD |

B

C

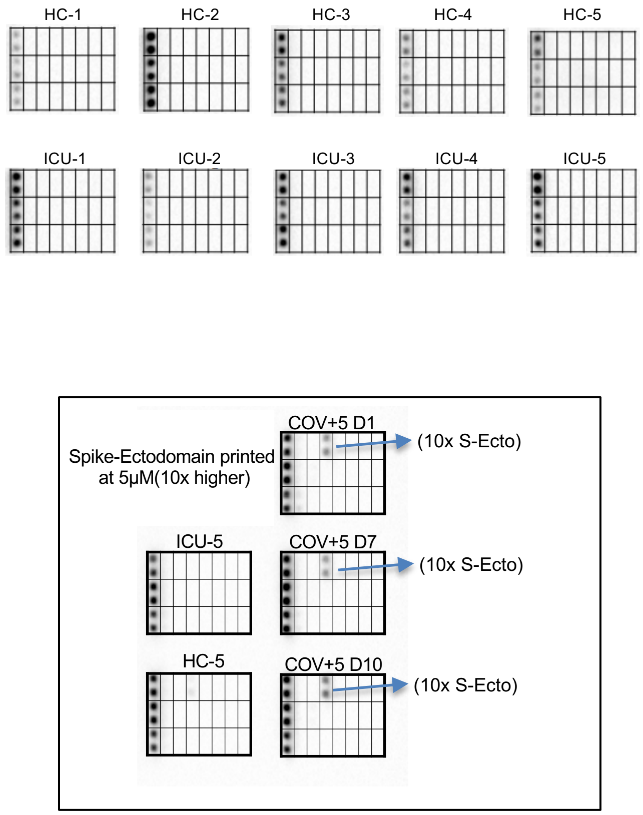


**Fig. S11. SARS-CoV-2 Proteome antigen array**. (A) Identity and position of proteins printed in the proteome array (see also Fig. 5). (B) The plasma from the ICU (COVID-19 negative) and healthy control group showed no antibody response to any virus antigen. (C) A proteome array printed with 10 x spike Ecto domain (5 μM) was screened with the plasma samples from the COV+5 patient
