## Supplementary material for "System-wide hematopoietic and immune signaling aberrations in COVID-19 revealed by deep proteome and phosphoproteome analysis": Table S1, S2 and supplementary text

### Table S1. Subject demographics and clinical data.

| Group | COVID+ patients | | | | | Non-COVID ICU patients | | | | |
|---|---|---|---|---|---|---|---|---|---|---|
| Sample ID | COV-1 | COV-2 | COV-3 | COV-4 | COV-5 | ICU-1 | ICU-2 | ICU-3 | ICU-4 | ICU-5 |
| Age range | 61-65 | 61-65 | 61-65 | 45-50 | 51-55 | 55-60 | 61-65 | 61-65 | 41-45 | 55-60 |
| Sex | M | F | F | M | M | M | F | F | M | M |
| Multiple organ dysfunction score | 8 | 4 | 11 | 5 | 5 | 5 | 0 | 1 | 3 | 7 |
| Sequential Organ Failure Assessment Score | 7 | 2 | 11 | 2 | 11 | 5 | 3 | 4 | 5 | n/a |
| Comorbidities | | | | | | | | | | |
| Hypertension | YES | NO | YES | NO | NO | YES | NO | NO | YES | NO |
| Diabetes | NO | NO | YES | NO | YES | NO | YES | NO | NO | NO |
| Chronic kidney disease | NO | NO | NO | NO | NO | NO | NO | NO | NO | NO |
| Cancer | NO | NO | NO | NO | NO | NO | NO | NO | NO | NO |
| Chronic obstructive pulmonary disease | NO | NO | NO | NO | NO | NO | YES | NO | NO | NO |
| Coronary artery/heart disease | NO | NO | NO | NO | YES | NO | YES | NO | NO | NO |
| Chronic heart failure | NO | NO | NO | NO | NO | NO | YES | NO | NO | NO |
| Baseline labs | | | | | | | | | | |
| WBC | 12.9 | 6.9 | 41.5 | 10.5 | 23.6 | 7.9 | 6.3 | 19.3 | 11.9 | 17.9 |
| Neutrophils | 11.1 | 5.8 | 24.1 | 9.7 | 21.1 | 7.2 | 5.3 | 16.1 | 10.6 | 15.6 |
| Lymphocytes* | 0.9 | 0.8 | 0.7 | 0.3 | 0.5 | 0.3 | 0.6 | 1.4 | 0.4 | 1.2 |
| Hemoglobin | 122 | 110 | 94 | 134 | 93 | 137 | 75 | 81 | 124 | 109 |
| Platelets | 186 | 203 | 366 | 216 | 463 | 291 | 261 | 163 | 248 | 259 |
| Creatinine | 993 | 56 | 565 | 78 | 82 | 45 | 56 | 80 | 46 | 71 |
| CXR finding | bilateral pneumonia | bilateral pneumonia | bilateral pneumonia | bilateral pneumonia | unilateral pneumonia | unilateral pneumonia | bilateral pneumonia | unilateral pneumonia | normal | unilateral pneumonia |
| Pao2/Fio2 ratio | 59 | 88 | 120 | 50 | 160 | 208 | 143 | 316 | 363 | n/a |
| Sepsis diagnosis | Confirmed | Confirmed | Confirmed | Confirmed | Confirmed | Suspected | Suspected | Suspected | Suspected | Suspected |
| Interventions during study | | | | | | | | | | |
| Antibiotics | YES | YES | YES | YES | YES | YES | YES | YES | YES | YES |
| Anti-virals | NO | NO | NO | NO | NO | NO | NO | NO | YES | YES |
| Steroids | NO | NO | NO | NO | YES | NO | YES | YES | NO | NO |
| Vasoactive medications | YES | YES | YES | YES | YES | NO | NO | YES | NO | YES |
| Renal replacement therapy | YES | NO | YES | NO | NO | NO | NO | NO | NO | NO |
| High-flow nasal cannula | NO | YES | NO | YES | NO | NO | NO | NO | NO | NO |
| Non-invasive mechanical ventilation | YES | NO | NO | NO | NO | YES | YES | YES | NO | NO |
| Invasive mechanical ventilation | YES | YES | YES | YES | YES | YES | NO | NO | YES | YES |
| Patient Outcome | DEAD | DEAD | DEAD | DEAD | ALIVE | ALIVE | ALIVE | ALIVE | DEAD | DEAD |

* 1.0-4.0 ($10^9$/L) for healthy adults

**Table S2. The list of samples for TMT labeling and mass spectrometry analysis.**

| Subject | Age range | Sex | Day collected | TMT batch & channels |
|---|---|---|---|---|
| COV1 | 61-65 | M | day1 | A9, C2 |
| | | | day7 | A2 |
| | | | day10 | A6 |
| COV2 | 61-65 | F | day1 | A10, C3 |
| | | | day7 | A4 |
| | | | day10 | A7 |
| COV3 | 61-65 | F | day1 | A11, C4 |
| | | | day7 | A5 |
| | | | day10 | A8 |
| COV4 | 45-50 | M | day1 | B2 |
| | | | day7 | B5 |
| | | | day10 | B7 |
| COV5 | 51-55 | M | day1 | B4 |
| | | | day7 | B6 |
| | | | day10 | B8 |
| ICU1 | 55-60 | M | day1 | B3, C5 |
| ICU2 | 61-65 | F | day1 | C6 |
| ICU3 | 61-65 | F | day1 | C7 |
| ICU4 | 41-45 | M | day1 | B9 |
| ICU5 | 55-60 | M | day1 | C8 |
| HC1 | 55-60 | M | | A3, C9 |
| HC2 | 61-65 | F | | C10 |
| HC3 | 61-65 | F | | C11 |
| HC4 | 46-50 | M | | B10 |
| HC5 | 51-55 | M | | B11 |
| PBMC, PV+ | | | | A1, B1, C1 |

Note: The channel 3 data (A3, B3 and C3) were not used for analysis due to strong reporter ion interferences from channel 1 (pervanadate-treated PBMC booster channel, A1, B1 and C1).

### SUPPLEMENTARY METHODS

#### Study design and blood sample collection

This study was approved by the Western University, Human Research Ethics Board (study number: 116284). Patients were admitted to level-3 academic intensive care unit (ICU) at London Health Sciences Centre-Victoria Campus (London, Ontario) and were suspected of having COVID-19 based on standard hospital screening procedures [1]. Blood samples were collected starting at admission for COVID-19$^{-}$ patients, or day 1, 7 and 10 for COVID-19$^{+}$ patients. COVID-19 status was confirmed as part of standard hospital testing by detection of two SARS-CoV-2 viral genes using polymerase chain reaction. Although ICU severity of illness scores have not been validated in COVID-19+ patients, we calculated multiple organ dysfunction score (MODS) and Sequential Organ Failure Assessment (SOFA) score for both COVID-19+[2].

Final participant groups were constructed by age- and sex-matching COVID-19$^{+}$ ICU patients with COVID-19$^{-}$ ICU patients, as well as healthy controls that had blood samples previously banked in the Translational Research Centre, London, ON, Canada) (directed by Dr. D. D. Fraser; https://translationalresearchcentre.com/)[3,4].

The peripheral blood mononuclear cell (PBMC)/buffy coat and plasma samples were de-identified prior to transfer from the hospital to a biosafety Level 3 (CL3) lab (ImPaKT, Western University) following Transportation of Dangerous Goods (TDG) guidelines. All plasma samples were heat-inactivated at 56 °C for 30 minutes and PBMCs were lysed in 9M Urea in HEPES buffer (20mM HEPES, 1mM sodium orthovanadate, 10mM NaF, pH8.0) at the ImPaKT CL3 facility as per Western university biosafety regulation. Heat inactivated plasma and lysed PBMCs samples were verified free of virus before they were transferred to the testing laboratory.

#### Sample processing for proteomics and phosphoproteomics analyses by mass spectrometry

##### *Hemoglobin depletion and protein precipitation*

Hemoglobin was depleted from PBMC whole cell lysate samples according to HemogloBind$^{TM}$ manufacturer instruction with modifications. Briefly, 10 ml HemogloBind beads were added to 1 ml whole cell lysate suspended in 20mM HEPES buffer pH 6.5 containing 1mM sodium orthovanadate, 10mM NaF, and the mixture was vortexed for 10 min at room temperature. The Mixture was then centrifuged for 5 minutes at 10,000 rpm and the protein supernatant was collected and precipitated with 5 volumes of ice-cold acetone/ethanol/acetic acid/ (v/v/v/=50/50/0.1) at -20 °C overnight. Protein pellets were collected by centrifugation at 17,000g for 20 min the following day and the resulting pellets were washed with ice-cold 75% ethanol once and centrifuged at 17,000g for 3min. Ethanol was removed and pellets were dried briefly and then resuspended in urea lysis buffer (9M urea, 20 mM HEPES, 1 mM sodium orthovanadate, 10mM NaF, pH8.0)

*Protein processing and digestion*

Protein concentration was estimated by Bio-Rad protein assay kit. The protein concentration was adjusted to 8 µg/µl in urea lysis buffer and reduced with 10 mM dithiothreitol (DTT) for 1 hour at room temperature. Protein was then alkylated with iodoacetamide (IAA) to a final concertation of 10 mM IAA followed by incubation for 45min in the dark at room temperature. Protein solution was then diluted 1:3 (vol/vol) with digestion buffer (50 mM HEPES, 1 mM orthovandate, 10 mM NaF, pH 8.0) to decrease urea concentration, LysC was then added in a ratio of 1 mAU per 50 µg of total protein followed by incubation of 2 hours at 25°C with gentle shaking. Trypsin was then added in a ratio 1:50, and incubated overnight at 28°C. The resulting peptide was desalted using SepPak C18 cartridges (Waters WAT054955) and SpeedVac dried.

*Pervanadate treatment for the booster channel PBMC*

PBMCs isolated from normal blood samples was treated by pervanadate as descried in [5] with modifications. Briefly, the pervanadate solution was prepared by adding 10 µl of 0.1 M sodium orthovanadate to 10 µl of 0.2 M hydrogen peroxide (diluted 50-fold from a 30% stock). The solution was then incubated at room temperature for 15 min. Excess hydrogen peroxide was inactivated by adding 2 µl of catalase in PBS (10 mg/ml). PBMCs were suspended in 5 volumes 0.1 mM pervanadate in a buffer containing 20 mM HEPES-NaOH (pH 7.4) and incubated at 37° for 10 min.

*Tandem Mass Tag (TMT) labelling*

For mass spectrometry analysis, we labelled 25 patient or healthy control samples with the 11-plex TMT isobaric labelling reagent (Thermo A37725) (Table S2) for sample identities with TMT set/channel numbers). In addition, we employed the pervanadate BOOST channel approach [6] by including the pervanadate-treated human PBMC sample as the reference channel (channel 1 of each 11-plex sample). Three sets of 11-plex reagents were used to label all samples.

The TMT labelling procedure was modified from [7]. The desalted peptides were reconstituted in 0.1% formic acid to determine peptide concentration by the BCA protein assay kit (Pierce 23225). 200 µg portions of peptides from each sample were aliquoted and vacuum-dried to evaporate formic acid. Each of 0.8 mg 11-plex TMT labelling reagents was reconstituted in 41 µl acetonitrile. The peptides were reconstituted in 40 µl of 50 mM HEPES (pH 8.5) to prepare 5 mg/ml peptide solution and were mixed with 20.5 µl of the TMT reagent to label the peptides at room temperature for two hours. The reaction was quenched by adding 4 µl of 5% hydroxylamine. The 11 samples were combined (2.2 mg total peptides) and desalted by the SepPak C18 cartridge.

We previously reported that superbinder SH2 domains could be used to enrich pTyr peptides for mass spectrometry analysis [8]. For enrichment of pTyr peptides, the SH2 superbinder was covalently coupled to agarose beads (Thermo 20402). The digested peptides were reconstituted in 50 mM ammonium bicarbonate and incubated with the SH2 superbinder beads for 30 min at room temperature with rotation. The flow-through fraction was saved for later analysis. The beads were washed four times with the same buffer. The bound pTyr peptides were eluted by 0.4% trifluoroacetic acid (TFA). The eluted peptides were loaded on the High pH peptide fractionation kit (Pierce 84868). The peptides were separated into eight fractions and the fractions were combined into four vials for mass spec injections.

The flow-through fraction contains peptides not captured by the superbinder SH2 domain. A 100 µg portion was separated into 12 fractions by the High-pH fractionation kit for full proteome analysis. A 500 µg portion was used for phosphopeptide enrichment by the $Ti^{4+}$-IMAC resin [9]. The phosphopeptides were eluted by 10% ammonia and vacuum-dried. The dried phosphopeptides were separated into eight fractions and then combined into four vials for mass spec injections.

*LC-MS/MS experiments*

The peptides were analyzed by the data-dependent acquisition method on the EASY-nLC 1000 system coupled to the Q-Exactive Plus mass spectrometer (Thermo Scientific). The peptides were separated on an EASY-Spray ES803A analytical column (Thermo Scientific) applying a flow rate of 300 nl/min and a linear gradient from 3 to 40% acetonitrile in 0.1% formic acid. The gradient length was 2 hours for pTyr phosphoproteome fractions, 4 hours for full proteome and IMAC phosphoproteome fractions. See Table S3 for mass spectrometry data acquisition parameters.

Peptide identification and quantification were performed using MaxQuant version 1.6.17.0 [10]. The mass spectra were searched against the human SwissProt sequences (20367 entries, downloaded on February 7, 2020), supplemented with common contaminants. For all searches, cysteine carbamidomethylation was set as the fixed modification, and methionine oxidation and N-terminal protein acetylation are set as variable modifications. Trypsin/P was specified as the proteolytic enzyme with up to two missed cleavage sites allowed. For IMAC- and superbinder (pTyr)-enriched datasets, Phospho (STY) was included as an additional variable modification. For phosphoproteomic data, the minimal peptide length for the search was set as 6, whereas the value was set as 7 for full proteome data. The multi-batch normalization option was used to normalize the three 11-plex datasets by selecting Channel 1 (common pervanadate-treated PBMC sample) as a reference channel, and the data were normalized by the weighted ratio to the reference channel. The reporter ion interference correction factors for the TMT reagent (Lot VB298423) were applied. The match-between-run option was turned on for peptide identification. Other MaxQuant parameters are left as default values.

*Proteome and phosphoproteome data analysis*

For data analysis, only the proteins (for proteome) or phosphosites (for phosphoproteome) observed in at least three samples in each of the five groups (COV-D1, D7, D10, ICU, HC) were retained. The median log2 value was subtracted from each sample to normalize the data. Phosphosites with the localization probability > 0.75 were retained. Perseus version 1.6.14.0 was used to analyze the data and to perform GO enrichment for pathways analysis [11]. For clustering and heatmap preparation of proteome and phosphoproteome data, the three groups (COV-D1, ICU and HC) were analyzed by the ANOVA test with 10% FDR cut-off, and the Z-score was calculated for each protein (for proteome) or phosphosite (for phosphoproteome) for hierarchical clustering. The data from PhosphositePlus version 110220 [12] were used to construct the kinase-pTyr substrate network figure in Figure 4. The list of the human kinases was based on [13]. The ITRM motifs were based on a list previously identified [14]. The KSEA App was used to predict active kinases [15].

Functional scores of phosphoprotein interactions were calculated by PHOTON (Rudolph et al., 2016). The Metascape server was used for gene annotation analysis [16]. The heatmaps were prepared with the Morpheus server. Cytoscape [17] was used to draw protein-protein interaction networks shown.

**RNA isolation and qPCR**

Isolation and purification of total RNA from the PBMCs was carried out according to RNeasy® Mini Kit followed by cDNA preparation using reverse transcriptase and random primers. The qPCR amplification was performed with cytokine/chemokine primers listed in the key resources table. After 40 cycles of PCR, ΔCt values were determined using different cytokine and chemokine primers and β-actin as reference genes. Differences in mRNA levels were then calculated using the 2−(ΔΔCt) method. The expression of human β-actin was used to normalize mRNA content and to calculate Log2 fold change in gene expression. Samples were tested five biological repeats and four technical repeats.

**Statistical analysis**

Statistical analyses were done using the GraphPad Prism9 software. Unpaired One-way ANOVA was conducted to test significance of difference of unpaired samples from different patient groups and repeated measure ANOVA was done for paired patients' samples as described in figure legends.

**Protein Microarray Printing**

SARS-CoV-2 proteins were diluted to 0.5-10µM in PBS with 5% glycerol (IgG control at 200nM) and aliquots transferred to a 384-well microplate (ArrayIt). 24 copies of the microarray were printed on each nitrocellulose coated glass slide (ArrayIt) using a VersArray Chipwriter Pro (Bio-Rad) equipped with a Stealth 15XB microarray quill pin (ArrayIt). Spot to spot distance was 850µm with two reprints of the same spot and all spots printed in duplicate in the y dimension. A dwell time of 0.1sec was used for each spot with an approach speed of 12.5mm/sec. Samples were printed at room temperature and subsequently stored at 4°C until time of probing.

For probing, microarray slides were briefly rinsed twice with TBST (Tris buffered saline containing Tween 20: 0.1M Tris-HCl, pH 7.4, 150mM NaCl, and 0.1% Tween 20) to wet the surface then incubated for 2h with ChonBlock ELISA blocking and antibody dilution buffer (Chondrex Inc). Slides were briefly rinsed with TBST then inserted into an ArraySlide 24-chamber hybridization cassette (The Gel Company) and incubated with plasma from SARS CoV-2 qPCR confirmed positive and negative patients (1:250 dilution in ChonBlock). Slides were then rinsed quickly three times followed by three 5min washes with TBST before probing with goat anti-human IgG HRP antibody at 1:10,000 (Millipore Sigma) in ChonBlock for 1h. The wash step was repeated as above, then the HRP signal was visualized on a ChemiDoc XRS+ Imager (Bio-Rad) using Clarity ECL Substrate (Bio-Rad). All incubation steps were performed at room temperature using a rocker for agitation of sample.

**Table of Key Regents**

| **REAGENT or RESOURCE** | **SOURCE** | **IDENTIFIER** |
|---|---|---|
| **Antibodies** | | |
| Anti-Spike-RBD antibody | Novus Biologicals | NBP2-90980 |
| Anti-nucleocapsid antibody | ThermoFisher Scientific | PA5-81794 |
| Goat anti-human IgG HRP antibody | Millipore Sigma | AP113P |
| Goat anti-rabbit IgG HRP | Bio-Rad | 170-6515 |
| **qPCR primers** | | |
| Human TGFβ<br>For: CAGCAACAATTCCTGGCGATA<br>Rev: AAGGCGAAAGCCCTCAATT | Millipore Sigma | |
| Human TNFα<br>For:<br>TCTTCTCGAACCCCGAGTGA Rev:<br>CCTCTGATGGCACCACCAG | Millipore Sigma | |
| Human IFNγ | Millipore | |

| | |
|---|---|
| For: TCAGCTCTGCATCGTTTTGG<br>Rev:<br>GTTCCATTATCCGCTACATCTGAA | Sigma |
| Human IL-8<br>For: GAACTGAGAGTGATTGAGAGT<br>Rev: CTTCTCCACAACCCTCTG | Millipore<br>Sigma |
| Human IL-6<br>For:<br>GTAGCCGCCCCACACAGACAGCC<br>Rev: GCCATCTTTGGAAGGTTC | Millipore<br>Sigma |
| Human IL-17<br>For: TGAAGGCAGGAATCACAAT<br>Rev: GGTGGATCGGTTGTAGTAAT | Millipore<br>Sigma |
| Human IL-13<br>For: ATTGCTCTCACTTGCCTT<br>Rev: GTCAGGTTGATGCTCCAT | Millipore<br>Sigma |
| Human IL-12<br>For: TGGAGTGCCAGGAGGACAGT<br>Rev: TCTTGGGTGGGTCAGGTTTG | Millipore<br>Sigma |
| Human IL-10<br>For: GTGATGCCCCAAGCTGAGA<br>Rev: CACGGCCTTGCTCTTGTTTT | Millipore<br>Sigma |
| Human IL-2<br>For:<br>AGAACTCAAACCTCTGGAGGAAG<br>Rev:<br>GCTGTCTCATCAGCATATTCACAC | Millipore<br>Sigma |
| Human IL-33<br>For: GCCTGTCAACAGCAGTCTACTG<br>Rev:<br>TGTGCTTAGAGAAGCAAGATACTC | Millipore<br>Sigma |
| Human IL-1a | Millipore<br>Sigma |

| | |
|---|---|
| For: TGTATGTGACTGCCCAAGATGAAG<br>Rev: AGAGGAGGTTGGTCTCACTACC | |
| Human IL-1b<br>For: CCACAGACCTTCCAGGAGAATG<br>Rev: GTGCAGTTCAGTGATCGTACAGG | Millipore Sigma |
| Human IL-15<br>For: AACAGAAGCCAACTGGGTGAATG<br>Rev: CTCCAAGAGAAAGCACTTCATTGC | Millipore Sigma |
| Human IL18<br>For: GATAGCCAGCCTAGAGGTATGG<br>Rev: CCTTGATGTTATCAGGAGGATTCA | Millipore Sigma |
| Human MIP-1b<br>For: GCTTCCTCGCAACTTTGTGGTAG<br>Rev: GGTCATACACGTACTCCTGGAC | Millipore Sigma |
| Human M-CSF<br>For: TGAGACACCTCTCCAGTTGCTG<br>Rev: GCAATCAGGCTTGGTCACCACA | Millipore Sigma |
| Human MCP-1<br>For: AGAATCACCAGCAGCAAGTGTCC<br>Rev: TCCTGAACCCACTTCTGCTTGG | Millipore Sigma |
| Human iNOS<br>For: GCTCTACACCTCCAATGTGACC<br>Rev: CTGCCGAGATTTGAGCCTCATG | Millipore Sigma |
| Human GB<br>For: CGACAGTACCATTGAGTTGTGCG<br>Rev: TTCGTCCATAGGAGACAATGCCC | Millipore Sigma |
| Human β-actin<br>For: CACCATTGGCAATGAGCGGTTC | Millipore Sigma |

| | | |
|---|---|---|
| Rev: AGGTCTTTGCGGATGTCCACGT | | |
| Human IL-4<br>For: TGCATTGTTAGCATCTCTTGA<br>Rev: CCCTTCTCCTGTGACCTCGTT | Millipore Sigma | |
| Human IFNa<br>For: GACTCCATCTTGGCTGTGA<br>Rev: TGATTTCTGCTCTGACAACCT | Millipore Sigma | |
| Human IFNb<br>For: CAACTTGCTTGGATTCCTACAAA<br>Rev: TATTCAAGCCTCCCATTCAATTG | Millipore Sigma | |
| ChonBlock ELISA blocking and antibody dilution buffer | Chondrex Inc | 9068 |
| Human IgG (200nM) | Equitech Bio inc | SLH66-0001 |
| Spike Receptor Binding Domain (RBD) (14µM) | ThermoFisher Scientific | RP-87678 |
| Spike Ectodomain (0.5µM) | (Hsieh et al. 2020) | ID: SARS-CoV-2 S HexaPro |
| NSP3-unique (10µM) | This Study | The Structural Genomics Consortium |
| NSP3-ADRP (10µM) | This Study | The Structural Genomics Consortium |
| NSP3-PLPro (10µM) | This Study | The Structural Genomics Consortium |
| NSP3-nucleic acid binding domain (10µM) | This Study | The Structural Genomics Consortium |
| NSP4CTD (10µM) | This Study | The Structural Genomics Consortium |
| NSP5 (10µM) | This Study | The Structural Genomics Consortium |
| NSP7 (10µM) | This Study | The Structural Genomics Consortium |
| NSP8 (10µM) | This Study | The Structural Genomics Consortium |
| NSP9 (10µM) | This Study | The Structural Genomics Consortium |
| NSP10 (10µM) | (Perveen et al. 2020) | The Structural Genomics Consortium |
| NSP16 (10µM) | (Perveen et al. 2020) | The Structural Genomics Consortium |
| Nucleocapsid (full length) (5µM) | RayBiotech | 230-01104 |
| Nucleocapsid dimerization domain (5µM) | This Study | The Structural Genomics Consortium |

| | | |
|---|---|---|
| Nucleocapsid RNA binding domain (5µM) | [18] | The Structural Genomics Consortium |
| **Critical Commercial Assays** | | |
| HemogloBind™ | Biotech Support Group | H0145-50 |
| RNeasy® Micro | Qiagen | Cat# 74004 |
| SensiFAST™ SYBR ® No-ROX Kit | | |
| Clarity ECL Substrate | Bio-Rad | 170-5060 |
| **Software** | | |
| MaxQuant | (Tyanova et al., 2016a) | 1.6.17.0 |
| Perseus | (Tyanova et al., 2016b) | v1.6.14.0 |
| Cytoscape | (Su et al., 2014) | v.3.8.2 |
| KSEA App | Wiredja et al. 2019 | v1.0 |
| Metascape | Zhou et al. 2019 | |
| Morpheus | | https://software.broadinstitute.org/morpheus/ |
| **Other** | | |
| SuperNitro microarray substrate slides | ArrayIt | SMN |
| Stealth 15XB microarray quill pin | ArrayIt | SMP15XB |
| Microarray Microplate 384 | ArrayIt | MMP384 |
| ArraySlide 24-4 hybridization chamber cassette | The Gel Company | AHS24-4 |
| Lysyl Endopeptidase (LysC) | Wako | 129-02541 |
| Trypsin | Promega | V5113 |
| TMT 11-plex isobaric label reagent | Thermo | A37725 |
| BCA protein assay kit | Pierce | 23225 |
| SulfoLink Coupling Resin | Thermo | 20402 |
| High pH Reversed-Phase Peptide Fractionation Kit | Pierce | 84868 |
| $Ti^{4+}$-IMAC resin | Dr. Mingliang Ye [19] | |
